# Tripping on context: Characteristics and predictors of placebo and nocebo psychedelic effects

**DOI:** 10.64898/2026.05.31.26354539

**Authors:** Madeline V. Stein, Matt Butler, Sarah Chapman, Quinton Deeley, Devin B. Terhune

## Abstract

Psychedelic drugs are emerging as potentially efficacious tools for treating psychiatric conditions and probing the neural basis of consciousness. Although drug administration context is widely believed to shape psychedelic effects, it remains unclear whether it can independently generate placebo and nocebo effects resembling psychedelic experiences and side effects. In a pre-registered experiment, 78 non-clinical participants inhaled inert medical air under placebo and control conditions while completing a time perception task and a resting-state period. In the placebo condition, the gas was presented as nitrous oxide, whereas in the control, it was correctly identified. Placebo administration increased altered states of consciousness, ego dissolution, dissociation, and side effects, but did not significantly impact time perception. Predictive modelling indicated that placebo-induced psychedelic effects were predicted by trait responsiveness to verbal suggestion and absorption. These findings demonstrate that context alone can induce psychedelic effects, with implications for its causal role in psychedelic action.

## Introduction

After a fallow period on the fringes of medicine and consciousness science, psychedelics are increasingly being investigated as potential treatments for a range of mental health conditions and as tools for the experimental investigation of consciousness (Carhart-Harris & Goodwin, 2017; Erritzoe et al., 2026). Recent clinical trials have renewed interest in these compounds as potential treatments for different psychiatric conditions (Andersen et al., 2021; Luoma et al., 2020; Wongpaiboon et al., 2025). In parallel, psychedelics are increasingly used as experimental models to investigate neuroplasticity, large-scale brain network dynamics, and the neurophysiological bases of conscious experience (Timmermann et al., 2023).

Despite this growing clinical and scientific interest, the empirical assessment of psychedelic effects is complicated by the influence of extra-pharmacological factors commonly described under the rubric of set and setting (Leary et al., 1963; Pronovost-Morgan et al., 2025; Viljoen et al., 2025). *Set* refers to an individual’s mindset at the time of drug administration, including expectations (Butler et al., 2022; Rucker, 2024; Szigeti & Heifets, 2024), prior psychedelic experiences, and individual difference factors (Aday et al., 2021; Vizeli et al., 2024), whereas *setting* encompasses the physical, cultural, and social environment of drug administration (Hartogsohn, 2016). Although set and setting are widely acknowledged as salient contributors that shape phenomenological responses to psychedelics (Carhart-Harris et al., 2018), they are rarely treated as causal variables in their own right or systematically examined under controlled experimental conditions (Glennon et al., 2025; Golden et al., 2022; Stein & Terhune, 2025). Consequently, the causal role of set and setting factors (henceforth referred to as context) in inducing psychedelic experiences remains poorly understood. In particular, the extent to which contextual features can be reliably isolated and experimentally characterised within controlled psychedelic paradigms has not been systematically examined.

Placebo and nocebo effects are widely recognised as arising from contextual features (Colloca & Barsky, 2020) many of which overlap with those implicated in psychedelic experiences (Pronovost-Morgan et al., 2023; Stein & Terhune, 2025). Despite their relevance, placebo effects are not adequately accounted for in most research on psychedelics due to poor placebo-control conditions (Wen et al., 2024), and high levels of participant and experimenter unblinding (e.g., awareness of drug condition) (Hovmand et al., 2023; Lii et al., 2023). In a similar vein, nocebo effects remain comparatively underacknowledged (Bălăeţ, 2024; Browne et al., 2025) because adverse events (and side effects) are not well defined, investigated, or reported in psychedelic research (Breeksema et al., 2022; van Elk & Fried, 2023). Accordingly, understanding the magnitude of placebo and nocebo effects in psychedelic contexts is essential for disentangling the neurochemical and contextual mechanisms contributing to both their therapeutic and adverse effects (Burke & Blumberger, 2021).

Verbal suggestions are a primary vehicle through which placebo and nocebo effects can be reliably induced (Colloca & Miller, 2011; Rooney et al., 2023; Stein et al., 2025b; Wager & Atlas, 2015). Suggestions are communications capable of producing or modifying psychological processes, often framed such that the response will be experienced as something that *happens* to the individual rather than as a deliberate action (e.g., “this medication will reduce your symptoms”) (Stein et al., 2025, April). Outside the context of psychedelics, suggestions have been shown to reliably induce a variety of anomalous perceptual states that closely parallel psychedelic experiences (Corlett et al., 2025; Deeley et al., 2012; Deeley et al., 2013; Evans & Lynn, 2021; Lynn & Evans, 2017; Terhune & Smith, 2006). Consistent with these findings, uncontrolled pilot work suggests that social observation and contextual features of a naturalistic psychedelic environment can induce psychedelic-like experiences in a subset of participants (Olson et al., 2020).

Beyond contextual influences, links between individual difference variables underlying psychedelic responses and placebo/nocebo effects point to further mechanistic overlap (Stein & Terhune, 2025). Trait responsiveness to verbal suggestions (REVS; also known as suggestibility), and absorption (Jamieson, 2005; Tellegen & Atkinson, 1974; Terhune & Jamieson, 2021), a tendency to become deeply engaged in an experience and lose awareness of one’s surroundings, have been shown to predict psychedelic responses (Aday et al., 2021; Studerus et al., 2012; Szigeti et al., 2024). Critically, these variables have also been found to predict both placebo effects (Corsi & Colloca, 2017; De Pascalis et al., 2002; Owens & Menard, 2011; Parsons et al., 2021) and nocebo effects (Bräscher et al., 2020; Stein et al., 2025a), and have also been implicated in the emergence of anomalous experiences in response to contextual cues (Granqvist et al., 2005; van Elk, 2015). Similarly, childhood trauma has been associated with increased symptom reporting and adverse outcomes in psychedelic contexts (Evans et al., 2023; Olofsson et al., 2026; but see also Mehmood et al., 2026), consistent with broader evidence linking trauma to negative symptom perception (Lüönd et al., 2025). Relatedly, individual differences in somatosensory amplification may differentially predict placebo and nocebo effects (Kunkel et al., 2025) in psychedelic contexts. These diverse findings suggest that individual difference predictors of psychedelic responses may partly reflect variability in responsiveness to suggestion and context-driven placebo and nocebo effects.

This pre-registered experiment examined whether presentation of a psychedelic context alone could induce psychedelic effects (placebo) and side effects (nocebo). In order to induce these effects, our context manipulation mirrored key informational and procedural features of psychedelic experiments spanning recruitment, contraindication screening, informed consent, and instructions prior to dosing. Participants inhaled medical air that was either incorrectly presented as nitrous oxide (placebo condition) or correctly presented as medical air (control condition). A placebo-N₂O model was chosen because its rapid onset/offset, safety profile, and experimental tractability (Brake et al., 2025; Piazza et al., 2022) make it well suited for controlled investigation of contextual influences on psychedelic experiences. Moreover, N₂O produces phenomenological and neurophysiological effects comparable to those of classic and non-classic psychedelics (Dai et al., 2023; Zorumski et al., 2025), and the procedural complexity and contextual salience of gas inhalation may augment the magnitude of placebo effects (Hróbjartsson & Gøtzsche, 2010).

We expected that the placebo condition would increase psychedelic experiences and side effects relative to the control condition and would alter subjective time perception during a temporal reproduction task (Wittmann et al., 2007; Yanakieva et al., 2019). We further used predictive modelling to examine whether REVS and different facets of dissociation (i.e., absorption, depersonalization/derealization) predicted placebo and nocebo effect magnitude, and whether somatosensory amplification and childhood trauma contributed to nocebo effects.

## Methods

This experiment was approved by the King’s College London Research Ethics Committee, conducted in accordance with the Declaration of Helsinki, and was pre-registered on the Open Science Framework (https://osf.io/4ab7r). The data are freely available (https://osf.io/4ab7r/overview).

### Design

The present experiment employed a counterbalanced repeated-measures placebo-controlled design (see **Figure 1**). A repeated-measures design was used to enable precise within-participant estimation of placebo and nocebo effects (Enck et al., 2013).

**Figure 1.**
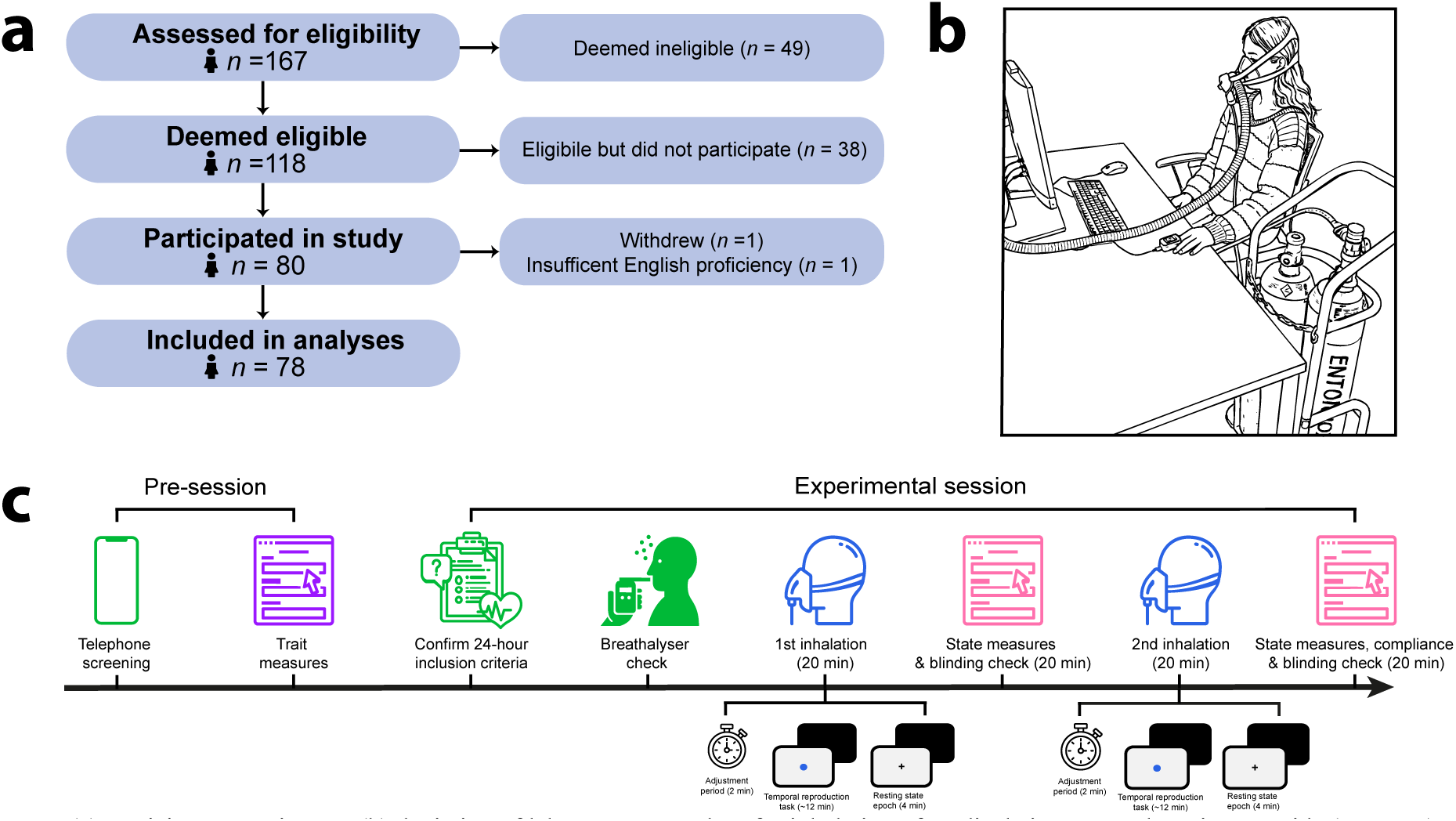
Experimental sequence and procedure of the placebo psychedelics experiment. ***Notes.*** (a) Participant recruitment; (b) depiction of laboratory procedure for inhalation of medical air presented as nitrous oxide (Entonox). The Entonox label was directly visible to participants and is only visually depicted in this manner to illustrate the label; and (c) Study timeline illustrating the sequence of procedures, beginning with contraindication screening and trait assessments, followed by the experimental session sequence.

### Participants

Our final sample included data from seventy-eight participants who were diverse in ethnicity and nationality (see **Supplementary Methods**), primarily female (41 females, 36 males, 1 undisclosed) and ranged in age from 18 to 44 (*M =* 25.23, *SD* = 6.20) with 0-12 years of post-secondary education (*M* = 4.11, *SD* = 3.32). The majority of participants were right-handed (94%) with normal or corrected-to-normal vision. A minority of participants had previously used nitrous oxide (N₂O), either within a medical context (16%) or recreationally (33%). Power analysis, exclusion details, and prior N_2_O use can be found in the **Supplementary Methods**.

### Materials

#### Trait measures

Participants completed a battery of baseline measures prior to the experiment including: REVS, assessed with the *Brief Suggestibility Scale* (BSS), which involves listening to a series of verbal suggestions followed by behavioural tests and self-report ratings of involuntariness (Wieder & Terhune, 2019); childhood trauma, assessed using the *Adverse Childhood Experiences* (ACE) Checklist (Felitti et al., 1998); dissociative absorption, depersonalization/derealization, and amnesia were measured with the *Dissociative Experiences Scale* (DES) (Carlson & Putnam, 1993); somatosensory amplification, assessed with the *Somatosensory Amplification Scale* (SAS) (Barsky et al., 1990); and compliant tendencies, measured with the *Gudjonsson Compliance Scale* (GCS) (Gudjonsson, 1989). We deviated from our pre-registration and omitted the *Acquiescence Response Set* (ARS) (Winkler et al., 1982), a measure of agreement with questionnaire statements irrespective of their content, from all analyses because it demonstrated poor internal consistency (⍺ = .17). All other measures and subscales demonstrated acceptable internal consistency in the present sample (⍺ range = .68-91; see **Supplementary Methods**).

#### State measures

Participants completed several state phenomenological measures indexing acute alterations in consciousness (i.e., placebo psychedelic effects), and side effects (i.e., nocebo psychedelic effects) following each inhalation condition. Shifts in perception, emotion, and self-experience commonly reported in psychedelic states were assessed across five dimensions using the *Altered States of Consciousness Questionnaire* (5D-ASC) (Dittrich, 1998); state dissociation including depersonalization, derealisation, and amnesia was assessed using the *Clinician Administered Dissociative States Scale* (CADSS) (Bremner et al., 1998); ego dissolution, reflecting positive and negative experiences of self-other boundary disintegration, was indexed using the *Ego-Dissolution Inventory* (EDI) (Nour et al., 2016); and physical and emotional side effects were assessed using the *General Assessment of Side Effects* (GASE) (Rief et al., 2011). All scales and subscales demonstrated good internal consistency in both conditions (⍺ range: placebo condition = .83-97; control condition =.70-96).

#### Temporal reproduction task

In each inhalation condition, participants completed three blocks of 30 trials of a temporal reproduction task, which has previously been shown to be sensitive to the influence of psychedelics (Wittmann et al., 2007; Yanakieva et al., 2019). Participants were instructed to estimate and memorise the duration of a blue circle, then reproduce the duration by holding down the space bar. Each trial began with a brief instruction cue (“memorise”; 750ms), followed by a blank, jittered inter-stimulus interval (425–650ms). Next, the target stimulus (blue circle: 80 × 80 pixels, ∼2 cm in diameter) displayed on a 1280 × 800 monitor appeared for a randomly varying interval (800 to 2600ms in 400ms steps). This was followed by a second blank inter-stimulus interval (500ms), after which a response cue (“reproduce”) signalled participants to reproduce the preceding target interval via a motor response. When participants pressed the space bar to begin reproducing the duration, the blue circle reappeared and remained on the screen until the space bar was released. A blank inter-trial interval (500ms) was presented before the next trial began.

#### Resting state epoch

At the end of each inhalation condition, participants completed a four-minute resting state epoch during which they fixated on a central cross on the computer monitor at a distance of ∼70-75cm.

#### Manipulation checks

Participants completed the *Guess of Treatment Questionnaire* (GoTq) (Szigeti et al., 2023), indicating whether they believed they had inhaled N₂O or medical air in each condition, alongside a single-item Likert measure assessing whether their responses were attributable to compliance (see **Supplementary Methods)**. Blinding was considered maintained when participants believed they had received N₂O during the placebo condition.

### Procedure

#### Recruitment and screening

Non-clinical participants were initially recruited through institutional recruitment sites, community-based flyers, social media, and word-of-mouth from the greater London area for an experiment investigating the impact of N_2_O on perception. To enhance the credibility of the advertised experiment’s aims, a trained screener administered a structured telephone screening to assess inclusion/exclusion criteria, similar to a genuine N_2_O experiment, including medical and psychiatric conditions, proneness to nausea, adverse reactions to N₂O, and frequent N₂O use (e.g., Das et al., 2016; Kamboj et al., 2020 and **Supplementary Methods**). Any uncertainty regarding eligibility was resolved in consultation with a psychiatrist (MB). In addition to confirming inclusion/exclusion criteria, the screener gave verbal suggestions for mild psychedelic effects (placebo) and side-effects (nocebo) pertaining to the subsequent experiment (see **Supplementary Methods**).

#### Experiment

In accordance with a genuine N_2_O experimental protocol (Das et al., 2016; Kamboj et al., 2020), participants were instructed to abstain from alcohol and drugs for 24 hours, avoid eating for four hours, and refrain from drinking fluids for two hours before the experiment for the purpose of reinforcing the credibility of the advertised experimental aims.

The in-person experiment took place at the Institute of Psychiatry, Psychology and Neuroscience at King’s College London. The experimenters included one or two women who ranged in age from 20-31 whereas the gas was administered by a third experimenter (a middle-aged white man); the team of experimenters was ethnically diverse and included individuals from different national backgrounds. The laboratory environment included a series of contextual cues to enhance the belief that participants would be receiving genuine N_2_O (**Figure 1**). We fitted a medical air cylinder with an *Entonox* label to mimic the widely-used agent (Piazza et al., 2022), which includes 50% N_2_O/50% O_2_. An inessential exhaust tube was routed from the inhalation mask to a window to ensure that any exhaled air was vented outside the testing room, thereby mimicking common ventilation procedures used in experimental N_2_O administration. The environment did not include plants, music, artwork, or other overt contextual cues (Pronovost-Morgan et al., 2025) and no other participants or confederates were present (Meeuwis et al., 2023; Olson et al., 2020).

Following confirmation of adherence to the 24-hour inclusion criteria via self-report and breathalyser testing (AlcoSAFE CA10 Breathalyzer; AlcoTech Incorporated), participants were instructed on how to signal their willingness to continue throughout the experiment and were fitted with a pulse oximeter (PO 80 Pulse Oximeter; Beurer) during inhalation periods. The use of physiological monitoring served as a contextual cue reinforcing the credibility of the experimental procedure and placebo manipulation; Oxygen saturation remained above 95% for all participants throughout the experiment.

The testing phase consisted of two 20-minute counterbalanced inhalation conditions. In the control condition, participants inhaled medical air correctly identified as such:

> *“In this part of the experiment, you will be inhaling Medical Air. As you read in the Information Sheet, this is similar to regular air. When inhaling medical air, many participants find that it feels just like they are breathing regular air in their day-to-day life. Medical air is completely safe.”*

By contrast, in the placebo condition, participants inhaled medical air incorrectly identified as Entonox:

> *“In this part of the experiment, you will be inhaling Entonox. As you read in the Information Sheet, this is a mixture of N_2_O and O_2_. When inhaling Entonox, many participants experience a variety of psychedelic effects including hallucinations, euphoria, changes in their perception of their body, and other types of anomalous experiences. However, bear in mind that these experiences are different for everyone, some people might have an intense experience while others might not. Entonox is completely safe, but some participants also experience minor side effects including nausea, dizziness, fatigue and other minor side effects.”*

Each 20-minute inhalation condition involved three phases (**Figure 1**): an adjustment period, completion of the temporal reproduction task, and a resting state epoch. After completion of inhalation, participants removed their masks and were given a 20-minute washout period, presented as an opportunity to eliminate any residual effects of the gas inhalation. During this period, participants completed the outcome measures in reference to the resting state epoch.

#### Analyses

All analyses were completed in *R* (R Core Team, 2023). The data analysis plan for this experiment was fully pre-registered (https://osf.io/4ab7r). Two deviations from the analysis plan were made due to unexpected violations of normality assumptions (see **Supplementary Analyses)**. Missing data for the ACE, SAS, GCS, and control EDI and CADSS were found for 0.2-4% of cases; all were judged to be missing completely at random based on a non-significant Little’s MCAR test (Little, 1988) and were imputed using the mice package (van Buuren & Groothuis-Oudshoorn, 2011). Three participants were excluded from relevant self-report analyses (see **Supplementary Analyses**), and five participants’ temporal reproduction data were excluded from relevant analyses due to technical issues resulting in lost data.

Paired-samples permutation *t*-tests (10,000 resamples) were used to compare phenomenological state measures across placebo and control conditions, with Hedges’ *g* and corresponding bootstrap (5,000 resamples) 95% confidence intervals [CIs] as a measure of effect size (Hedges, 1981). Hedges’ *g*s were coded such that positive values reflected a greater placebo/nocebo effect. For self-report measures, *t*-tests were supplemented with permutation 2 (Condition: control v. placebo) × 2 (Order: placebo-first v. placebo-second) ANOVAs to assess order effects. Significant interactions were decomposed using subsidiary permutation *t*-tests.

Temporal reproduction data were analysed using two complementary analyses (Yanakieva et al., 2019). First, we computed within-participant regression slopes at the individual participant level for each inhalation condition using all trial data by regressing reproduction times on stimulus intervals using robust regression. Individual regression slopes indexed the correspondence between stimulus and reproduced durations within each condition. In addition, median reproduction times for each participant were analysed using a 2 (Condition) × 10 (Interval) repeated-measures ANOVA including Greenhouse–Geisser corrections for all Interval effects and η_p_² as the effect size estimate.

Spearman correlations (ρ) were used to assess associations between variables. Predictive modelling was implemented through a series of regularized regression analyses to evaluate trait predictors of absolute difference scores (placebo-control) (Mattes & Roheger, 2020) across self-report outcomes, comparing models with ridge (α = 0), elastic net (α = .01–.99), and lasso (α = 1) penalties using the glmnet package (Friedman et al., 2010). The optimal mixing parameter (α) was selected based on test-set performance (see **Supplementary Table 3**), and the regularization parameter (λ) was determined via 10-fold cross-validation, using mean squared error (MSE) as the tuning criterion (Zou & Hastie, 2005).

To control for multiple comparisons, *p*-values from the permutation *t*-tests, repeated-measures ANOVAs, and correlation analyses were corrected using a False Discovery Rate (FDR) correction (Benjamini & Yekutieli, 2001).

## Results

### Magnitude of placebo and nocebo psychedelic effects

As shown in **Figure 2** and **Table 1**, the placebo condition was associated with significantly higher scores across all phenomenological state measures. A large effect was observed for depersonalisation (CADSS Dep), *g* = 0.60. Moderate effects (*g* range = 0.30–0.54) were observed for altered states of consciousness (5D-ASC), state dissociation (CADSS) and its remaining subscales, ego dissolution (EDI), and side effect severity (GASE). A weak effect was observed for dread of ego dissolution (DED), *g* = 0.19. Overall, these findings indicate that both placebo and nocebo effects were present across measures with effects that tended to be moderate in magnitude.

**Figure 2.**
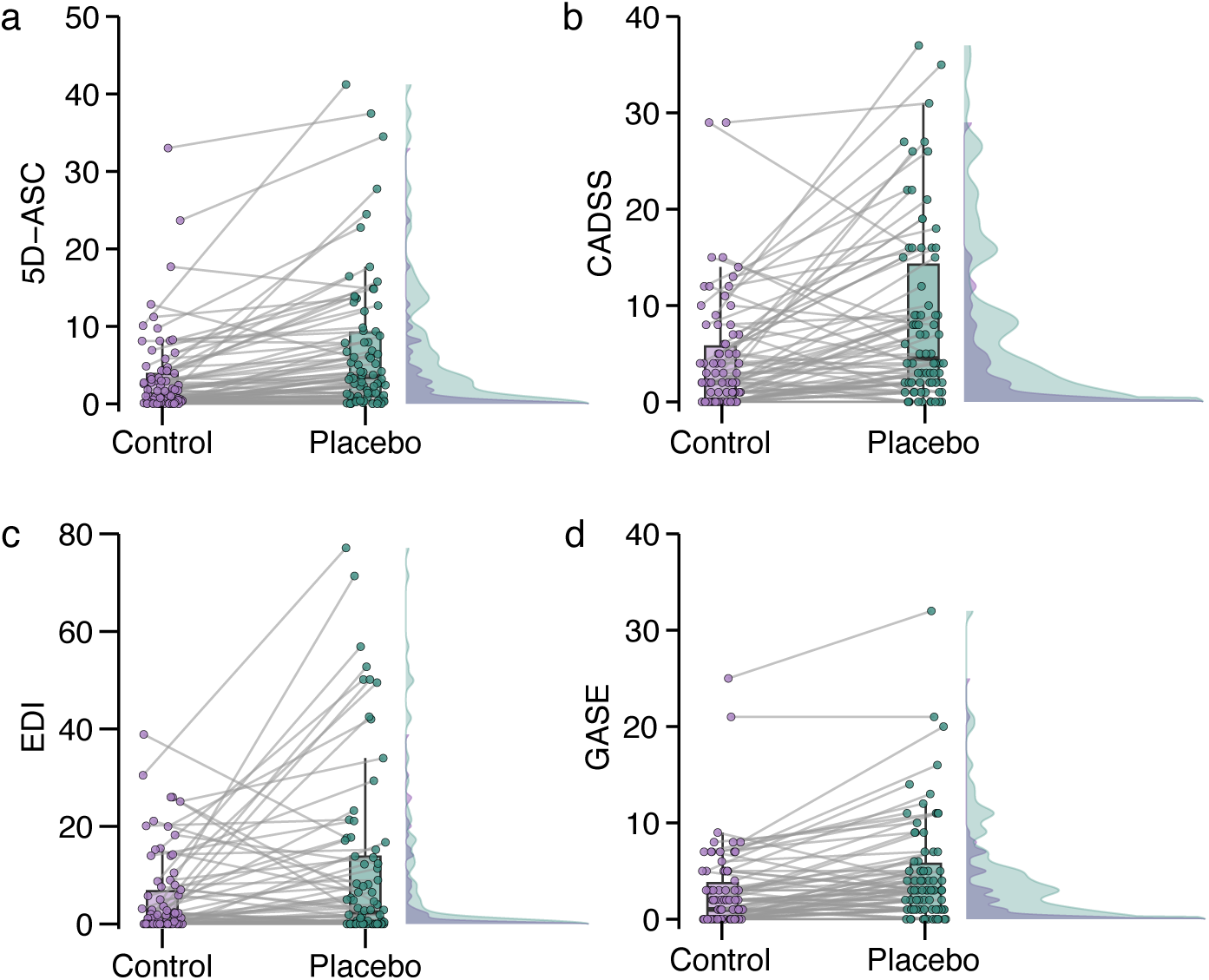
Raincloud plots showing psychedelic state and side effect measures across control and placebo conditions (N = 78). **Notes.** 5D-ASC = *5-Dimensional Altered States of Consciousness Rating Scale*; CADSS = *Clinician-Administered Dissociative States Scale*; EDI = *Ego Dissolution Inventory*; GASE = *Generic Assessment of Side Effects*.

**Table 1.** Descriptive and inferential statistics for outcome measures as a function of experimental condition (N = 78).

| Outcome | Control | Placebo | *t*(77) | *p* | *g* [95% CIs] |
| --- | --- | --- | --- | --- | --- |
|  | *M* (*SD*) | *M* (*SD*) |  |  |  |
| 5D-ASC | 3.26 (5.41) | 6.83 (8.76) | 5.35 | <.001 | 0.49 [0.33, 0.74] |
| 5D-ASC AUA | 1.24 (3.65) | 2.94 (7.50) | 3.31 | <.001 | 0.30 [0.20, 0.48] |
| 5D-ASC DED | 3.03 (8.79) | 5.00 (11.42) | 2.04 | .007 | 0.19 [0.04, 0.43] |
| 5D-ASC OBN | 1.72 (3.71) | 5.56 (11.34) | 3.28 | <.001 | 0.50 [0.34, 0.71] |
| 5D-ASC VIR | 12.32 (15.78) | 22.17 (20.47) | 5.49 | <.001 | 0.53 [0.35, 0.76] |
| 5D-ASC VRS | 1.61 (3.38) | 4.13 (7.59) | 3.84 | <.001 | 0.45 [0.29, 0.62] |
| CADSS | 4.26 (5.80) | 8.38 (9.18) | 4.75 | <.001 | 0.54 [0.32, 0.79] |
| CADSS Amn | 0.74 (0.99) | 1.14 (1.17) | 3.07 | .003 | 0.36 [0.13, 0.64] |
| CADSS Dep | 0.73 (1.34) | 2.09 (3.11) | 4.24 | <.001 | 0.60 [0.36, 0.84] |
| CADSS Der | 2.62 (3.81) | 5.15 (5.62) | 4.65 | <.001 | 0.53 [0.30, 0.79] |
| EDI | 5.18 (8.53) | 11.06 (17.94) | 3.21 | <.001 | 0.44 [0.21, 0.65] |
| GASE | 2.78 (4.20) | 4.72 (5.55) | 5.61 | <.001 | 0.39 [0.25, 0.63] |

**Notes.** 5D-ASC = 5-Dimensional Altered States of Consciousness Rating Scale; 5D-ASC subscales: AUA = Auditory Alterations; DED = Dread of Ego Dissolution; OBN = Oceanic Boundlessness; VIR = Vigilance Reduction; VRS = Visionary Restructuralization; CADSS = Clinician-Administered Dissociative States Scale; CADSS Amn = CADSS-Amnesia subscale; CADSS-Der = CADSS Depersonalization subscale; CADSS Der = CADSS-Derealization subscale; EDI = Ego Dissolution Inventory; GASE = Generic Assessment of Side Effect. An FDR correction yielded a corrected threshold of $p = .008$ . Means and SDs are reported for consistency with the primary permutation-based analyses.

We next assessed whether these effects were moderated by administration order, using permutation-based 2 (Inhalation condition: placebo v. control) × 2 (Order: placebo-first v. placebo-second) ANOVAs (see **Supplementary Table 2**). Significant Condition × Order interactions emerged for four of the 12 measures (EDI, CADSS, CADSS Dep, and CADSS Der subscales). In each case, decompositions revealed significant placebo effects in the placebo-first group that were large in magnitude (*g* range = 0.60–0.77), whereas effects in the control-first groups were non-significant and small in magnitude. These results suggest that placebo effects for approximately one third of the measured outcomes tended to be mostly specific to when the placebo condition preceded the control condition, implying an attenuation of placebo effects with greater familiarity with medical air inhalation.

### Temporal reproduction

Temporal reproduction task performance did not reliably differ between conditions (see **Supplementary Figure 3**). Within-participant regression slopes relating stimulus intervals to reproduction times did not significantly differ between placebo and control conditions, *t*(73) = 0.37, *p* = .71, *g* = 0.04 [−0.21, 0.26]. Similarly, a Condition × Interval repeated-measures ANOVA on reproduction times revealed no significant effects of Condition, *F*(1, 73) = 0.93, *p* = .34, η_p_² = .01, or a Condition × Interval interaction, *F*(6.21, 453.67) = 0.45, *p* = .85, η_p_² = .01, with near-zero effect sizes. As expected, there was a strong main effect of Interval, *F*(4.00, 291.75) = 350.59, *p* < .001, η_p_² = .82. These results indicate that reproduction times increased proportionally with stimulus interval in both conditions, indicating appropriate task performance, but did not significantly differ between placebo and control conditions.

### Assessment of blinding integrity

Participants’ confidence in their inhalation condition guess was moderate to high within both placebo and control conditions, and did not significantly differ between them, *Z* = 0.99, *p* = .32 (see **Supplementary Tables 2 & 3** and **Supplementary Figure 1)**. In the placebo condition, 45% of participants (*n* = 35) believed they received N_2_O, failing to correctly identify the placebo; among these, 44% received the placebo first. One participant in the blinded group gave a confidence rating of zero (equivalent to a random guess) and was removed from further blinding-related analyses. Among those who were blinded, participants reported significantly greater attribution of their condition guess to side effects and perceptual effects than to health improvements, *Z* = 4.27, *p* < .001. Of those who correctly identified the placebo and were thus unblinded (*n* = 43; 55% of total sample), 52% received the placebo first. Among the unblinded participants, condition guesses were significantly more strongly attributed to the lack of side effects than to the lack of health improvements, *Z* = 3.92, *p* < .001. The magnitudes of placebo and nocebo effects were not significantly larger in participants who remained blinded than those who were unblinded after correcting for multiple comparisons (**Supplementary Table 4**), with most effects remaining significant or near-significant in the latter group, although effects were reliably numerically larger in those who remained blinded. There were no significant trait predictors of blinding status (see **Supplementary Results**). These results indicate that just under half of participants were blind to the procedure’s status as a placebo, and blinding maintenance or loss was mostly due to the presence or absence of perceptual effects and side effects, although multiple weak-to-moderate placebo/nocebo effects were still observed in unblinded participants.

### Trait predictors of placebo and nocebo effects

Our next series of analyses assessed associations among trait variables and placebo and nocebo effects (see **Figure 3**). Difference scores (placebo-control condition) for state measures tended to be moderately-to-strongly intercorrelated although only altered states of consciousness (Δ5D-ASC) and its subscales were related to side effects (ΔGASE). Trait dissociation (DES) and compliance (GCS) were the only trait measures that correlated with placebo effects and significant trait-state correlations tended to be weak in magnitude (range: .29 to .38). Trait dissociation (DES and DES-absorption) correlated with increases in altered states of consciousness (Δ5D-ASC, Δ5D-ASC AUA, Δ5D-ASC OBN) and ego dissolution (ΔEDI) in the placebo condition; by contrast, trait depersonalisation (DES-DD) was only associated with Δ5D-ASC AUA. GCS scores were significantly associated with state dissociation effects (ΔCADSS, ΔCADSS-Dep, ΔCADSS Der), and ΔEDI whereas dissociative absorption (DES-absorption) was the only trait variable related to side effects (ΔGASE). Cumulatively, these findings suggest that trait-level predictors of placebo and nocebo effects are modest in magnitude, with dissociation and compliance emerging as the most consistent correlates.

**Figure 3.**
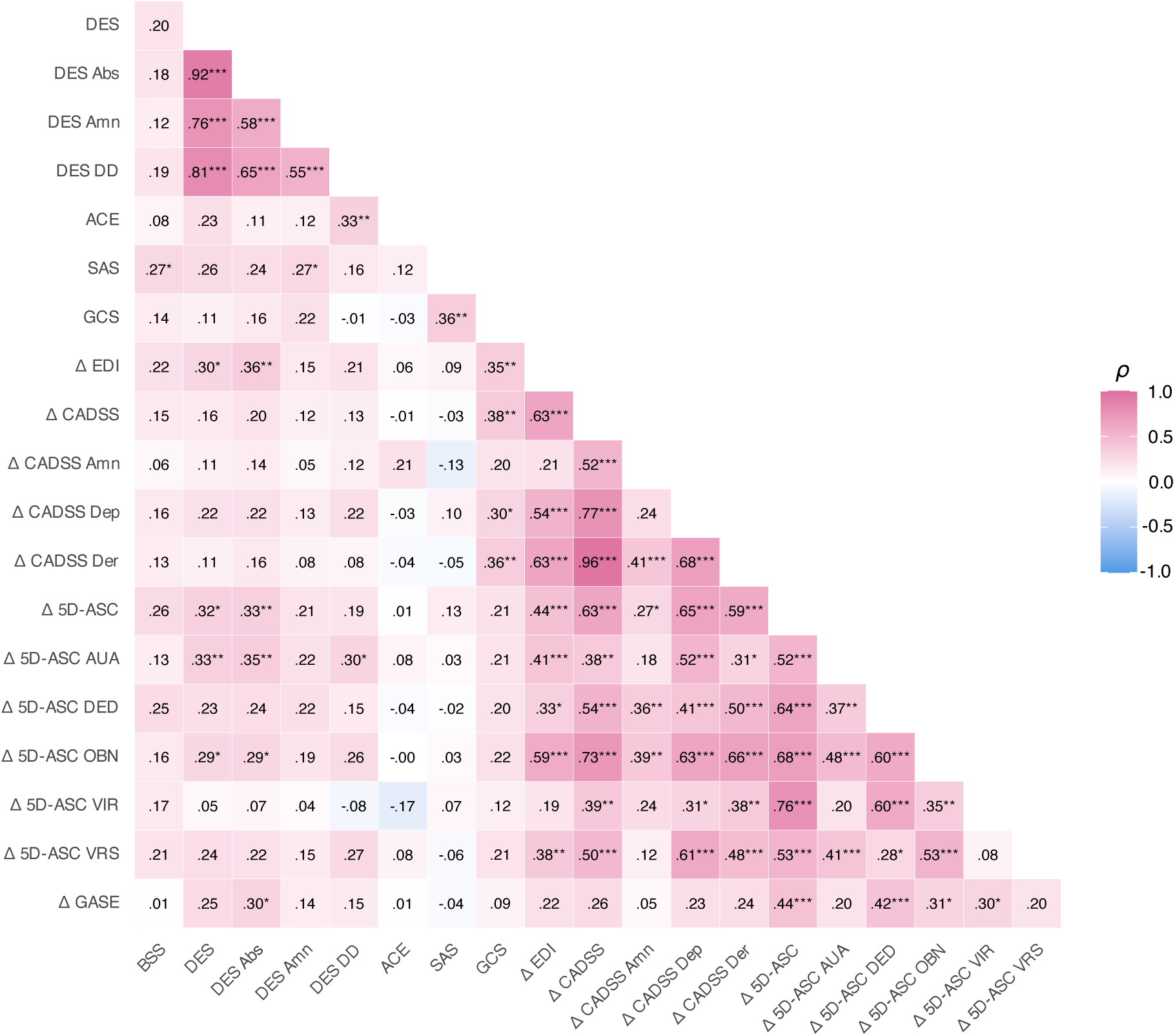
Heatmap of Spearman correlation coefficients between trait predictor variables and placebo and nocebo effect outcomes. Notes. Δ = absolute difference (placebo-control); ACE = *Adverse Childhood Experiences Checklist*; BSS = *Brief Suggestibility Scale*; CADSS = *Clinician-Administered Dissociative States Scale*; CADSS Amn = CADSS-Amnesia subscale; CADSS-Dep = CADSS Depersonalization subscale; CADSS Der = CADSS-Derealization subscale; EDI = *Ego Dissolution Inventory*; DES = *Dissociative Experiences Scale*; DES Abs = DES Absorption subscale; DES Amn = DES Amnesia subscale; DES DD = DES Depersonalization/Derealization subscale; GCS = *Gudjonsson Compliance Scale*; SAS = *Somatosensory Amplification Scale*; 5D-ASC = *5-Dimensional Altered States of Consciousness Rating Scale*; 5D-ASC subscales: AUA = Auditory Alterations; DED = Dread of Ego Dissolution; OBN = Oceanic Boundlessness; VIR = Vigilance Reduction; VRS = Visionary Restructuralization; GASE = *Generic Assessment of Side Effects*. * *p*<.044 (false discovery rate [FDR] threshold) ** *p*<.01 *** *p*<.001

Our final series of analyses used regularization regression to assess whether trait measures could collectively predict placebo and nocebo effects. A model of placebo-induced altered states of consciousness (Δ5D-ASC) accounted for a modest proportion of in-sample variance (*R*² = .11) with slightly lower cross-validated generalizability (CV *R*² = .07), with REVS, dissociation, dissociative absorption, and compliance identified as positive but weak predictors; REVS showed the greatest predictive efficacy (**Figure 4**; **Supplementary Table 5**). Similar patterns were observed for the 5D-ASC subscales of auditory alterations and oceanic boundlessness, whereas models for dread of ego dissolution, vigilance reduction, and visionary restructuralization yielded no significant predictors. Models predicting placebo-induced dissociation (ΔCADSS) accounted for a modest proportion of in-sample variance (*R*² = .15) albeit with poor cross-validated generalizability (CV *R*² = −.03) with a similar pattern for models predicting ΔEDI (*R*² = .12; CV *R*² = −.06). Finally, the model predicting side effects (ΔGASE) explained a modest proportion of in-sample variance (*R*² = .13) with weak cross-validated performance (CV *R*² = .05). Across these models, compliance and trait dissociation were consistently retained as predictors, although their effects were generally small (see **Supplementary Table 5** for full model details). Collectively, these models point to REVS as a specific predictor of altered states of consciousness, trait dissociation and dissociative absorption as more consistent predictors of a range of placebo psychedelic effects, and compliance as a weak predictor of dissociative responses, with limited evidence for reliable trait predictors of nocebo effects.

**Figure 4.**
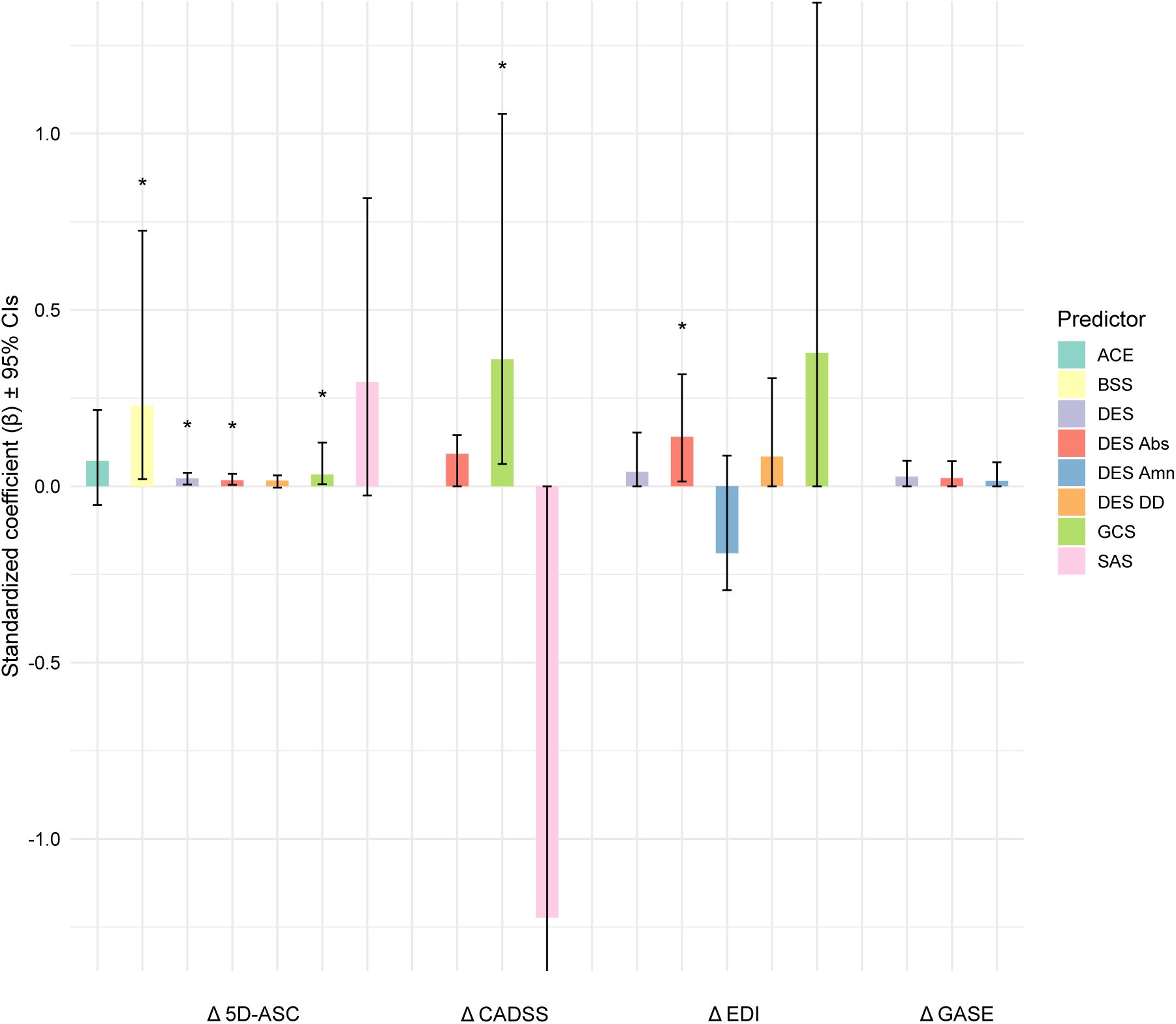
Regularized regression beta coefficients (with 95% confidence intervals) for trait predictors of placebo and nocebo effects. **Notes.** Δ = absolute difference (placebo-control); Outcomes: 5D-ASC = *5-Dimensional Altered States of Consciousness Rating Scale*; CADSS = *Clinician-Administered Dissociative States Scale*; CADSS Amn = CADSS-Amnesia subscale; CADSS-Dep = CADSS Depersonalization subscale; CADSS Der = CADSS-Derealization subscale; Predictors: ACE = *Adverse Childhood Experiences Checklist*; BSS = *Brief Suggestibility Scale*; EDI = *Ego Dissolution Inventory*; DES = *Dissociative Experiences Scal*e; DES Abs = DES Absorption subscale; DES Amn = DES Amnesia subscale; DES DD = DES Depersonalization/Derealization subscale; GCS = *Gudjonsson Compliance Scale*; SAS = *Somatosensory Amplification Scale*.

## Discussion

In a controlled laboratory experiment, we evaluated whether a psychedelic context alone could reproduce classic psychedelic effects and side effects. In line with a wealth of evidence demonstrating the efficacy of placebo and nocebo interventions (Colloca, 2024; Colloca & Barsky, 2020), we found that an inert procedure mimicking a psychedelic experiment reliably elicited psychedelic responses and side effects on standardised psychometric measures. Predictive modelling further demonstrated that placebo-induced psychedelic effects were associated with baseline psychological traits previously linked to outcomes in psychedelic research (Aday et al., 2021; Stein & Terhune, 2025; Szigeti et al., 2024). These results demonstrate that a psychedelic context alone, without pharmacological action, can partially reproduce psychedelic effects, positioning context itself as a causal contributor to the emergence of psychedelic experiences and a salient target for clinical and experimental research on psychedelics.

Our central result was that a placebo intervention comprising the administration of medical air presented as N_2_O and accompanying verbal suggestions yielded both psychedelic effects and side effects. We induced these effects by mirroring the informational context of a psychedelic experiment, introducing verbal and non-verbal suggestions for psychedelic effects throughout multiple stages of the experiment spanning recruitment, contraindication screening, informed consent, and instructions prior to dosing. The observed placebo and nocebo effects tended to be moderate in magnitude, consistent with meta-analyses of these effects in other contexts (Blythe et al., 2023; Peerdeman et al., 2016; Rooney et al., 2024; Stein et al., 2025b). The largest placebo effects were observed with measures of depersonalisation and derealisation, followed by indices of ego dissolution, altered states of consciousness and visual imagery, with the weakest effects observed with dissociative amnesia, auditory alterations and dread of ego dissolution. Notably, these effects were observed on widely used measures within psychedelic research (Brake et al., 2025; Hovmand et al., 2024), underscoring the extent to which core features of the psychedelic experience can be reproduced in the absence of pharmacological action. These effects broadly map onto our verbal suggestions although the greater occurrence of dissociative responses may be attributable to participants’ awareness of N_2_O as a dissociative anaesthetic (Kronenberg et al., 2024), alongside broader cultural associations between N₂O (“laughing gas”), altered states, and psychedelics more broadly. Conversely, no significant placebo effects were observed on temporal reproduction performance, a domain outside the scope of the suggested phenomenology.

A salient result of this experiment was that our placebo procedure elicited reliable nocebo effects. This result align with broader calls for the systematic identification and assessment of adverse events and side effects within psychedelic studies (Taillefer de Laportalière et al., 2023; van Elk & Fried, 2023). As critiques of psychedelic research have increasingly focused on placebo effects (Butler et al., 2022; Szigeti & Heifets, 2024), comparatively less attention has been afforded to nocebo processes (Bălăeţ, 2024; Browne et al., 2025) and their role in generating side effects in psychedelic contexts. The moderate magnitude of nocebo effects observed in the present experiment suggests that side effects in psychedelic research are not solely pharmacologically driven but are at least partly shaped by contextual factors, with important implications for both research and clinical practice insofar as side effects may be systematically generated by features of the experimental or therapeutic context (Stein & Terhune, 2025). It also has bearing on the role of side effects in psychedelic experiences as drugs with more potent effects have been shown to elicit larger placebo effects (e.g., Schenk et al., 2024; Wiech et al., 2024). Greater attention to the communication of potential side effects in psychedelic trials as well as the identification, measurement, and modulation of nocebo processes is necessary to more accurately characterise risk and optimise outcomes in psychedelic interventions.

Although the placebo procedure reliably elicited psychedelic effects and side effects with moderate effect sizes, features of our experimental design likely attenuated the magnitude of these responses. Although a repeated-measures cross-over design was necessary to precisely quantify placebo and nocebo effects at the individual participant level (Enck et al., 2013) to enable predictive modelling of outcomes at the group level (see below), this design seems to have reduced the aggregate placebo effect. In particular, placebo effects tended to be larger in participants who completed the placebo condition prior to the control condition relative to those who received the control first. A weaker placebo effect in the latter group plausibly reflects increased familiarity with medical air following initial exposure, facilitating its detection in the subsequent placebo condition and/or a recalibration of expectations (Mestre, 2020). Alternatively, contrast effects or counter-conditioning may have contributed, whereby initial exposure to an inert condition establishes a conservative reference point that attenuates subsequent responses (Bieniek & Bąbel, 2023; Rubanets et al., 2026). In addition, placebo and nocebo effects tended to be numerically, albeit not statistically, larger in participants who remained blinded to the placebo procedure, implying that unblinding may have attenuated effect sizes (Aday et al., 2025) and that placebo effects will be larger as participant blinding rates are enhanced. Finally, our measurement of the nocebo effect likely reflects an underestimate as we used a general measure of side effects (Rief et al., 2011) that indexed symptoms that were irrelevant to, or implausible within, the experimental context and specific suggestions. Collectively, these experimental features arguably attenuated aggregate placebo and nocebo effects and suggest that placebo-induced psychedelic effects are substantially larger – and possibly account for a greater proportion of psychedelic outcome effects – than the present results imply.

Individual difference variables relevant to psychedelic contexts (Aday et al., 2021; Stein & Terhune, 2025) partly accounted for variability in placebo, but not nocebo, psychedelic effects. Trait REVS was an independent predictor of placebo psychedelic effects on the 5D-ASC but no other outcomes, which is consistent with previous research showing that REVS predicts antidepressant response to psychedelics (Szigeti et al., 2024) as well as response to placebo procedures (De Pascalis et al., 2001; Parsons et al., 2021). The specificity of this association is potentially attributable to our experimental suggestions (Bschor et al., 2024; Weng et al., 2025), which targeted perceptual effects assessed by the 5D-ASC (Dittrich, 1998). By contrast, trait dissociative absorption was a key predictor of placebo psychedelic effects across multiple outcomes and was the only significant correlate of the nocebo effect although it was not retained in our regression model. This aligns with research demonstrating that trait absorption predicts induced state dissociation (Condon & Lynn, 2014), psychedelic responding (Studerus et al., 2012) and nocebo responding and heightened symptom perception (Bräscher et al., 2020; Stein et al., 2023). Rather than reflecting trait sensitivity to psychedelic effects (Studerus et al., 2012), these diverse data suggest that absorption reflects a proneness to anomalous experiences (Glicksohn & Barrett, 2003; Terhune & Jamieson, 2021) or a trait-like perceptual sensitivity to context (Granqvist et al., 2005). Compliance also emerged as a significant predictor of dissociative responses. Given the extensive reliance on self-report measures in psychedelic research (see Prugger et al., 2022), this raises the possibility that some psychedelic effects may be inflated by compliant responding. Cumulatively, these results indicate that placebo psychedelic effects are at least partially attributable to individual differences in REVS, dissociation, and compliance, underscoring the importance of systematically assessing these constructs within this research domain.

Alongside individual difference factors, the present findings highlight methodological features as an additional source of variability in psychedelic research. Order effects were observed for state dissociation and ego dissolution, with higher scores among participants who received the placebo prior to the control condition. These findings point to the importance of rigorous counterbalancing, adequate washout periods, and explicit modelling of order in statistical analyses (Aday et al., 2022; Colloca & Fava, 2024). Examination of participants’ attribution patterns indicated that blinding guesses were informed by the experience of perceptual effects and side effects, with blinded participants more frequently attributing their guesses to such effects, whereas unblinded participants tended to attribute their guesses to the absence of such effects. This pattern is consistent with the content of the experimental suggestions, which emphasised side effects but did not reference health improvements, suggesting that blinding was primarily maintained through responsiveness to the experiment’s suggestions. Further well-powered research is needed to identify the predictors of blinding status in both placebo and pharmacological psychedelic research.

### Strengths & limitations

Among the strengths of the present experiment are the use of pre-registered controlled experimental design; a relatively large and ethnically diverse sample; the use of a complex placebo procedure, as more invasive placebos have been shown to produce larger placebo effects (Hróbjartsson & Gøtzsche, 2010; Kaptchuk et al., 2000; Kaptchuk et al., 2008; Meissner et al., 2013); and standardised experimental communications with carefully matched placebo and control conditions. Nonetheless, the findings of this experiment should be interpreted in light of multiple limitations. The use of a highly complex placebo induction procedure may limit the generalisability of these findings to more common psychedelic administration modalities (e.g., oral dosing), as most studies do not involve gas inhalation and mode of administration moderates the magnitude of both placebo (Benedetti & Dogue, 2015) and nocebo effects (Rooney et al., 2024; Stein et al., 2025b). Although the total sample provided sufficient statistical power to detect placebo and nocebo effects and their predictors, subgroup comparisons and trait predictors of blinding status were likely underpowered. Finally, our findings primarily rely on self-report measures, which may be subject to bias, although the methodological inferiority of such measures is contested (Corneille & Gawronski, 2024). Moreover, reliance on such measures reflects a broader limitation within psychedelic research, where there is no consensus on behavioural markers (Aday et al., 2022; Hovmand et al., 2024), and perceptual effects are widely considered the primary outcome of interest of direct relevance to clinical outcomes (Bhatt et al., 2025; Yaden & Griffiths, 2021).

### Implications

These results necessitate a revision of the theorised role of context in psychedelic experiences. Although psychedelic research has long recognised that experiences are shaped by the administration context (Carhart-Harris et al., 2018), this framework treats context as a secondary, moderating factor that is inseparable from pharmacological action (Pronovost-Morgan et al., 2023). In parallel, placebo effects have largely been viewed as a nuisance variable within psychedelic research. By demonstrating that a psychedelic context alone can elicit both psychedelic effects and side effects, the present findings challenge these assumptions and indicate that context can function as an independent causal driver of these outcomes. This aligns with broader shifts away from the conceptualisation of placebo effects as a variable to be controlled to an important neurobiological source of variability in response to active interventions (Ashar et al., 2017; Burke et al., 2026; Colloca & Barsky, 2020; Wager & Atlas, 2015). This has direct implications for both experimental and clinical practice: effects attributed to psychedelics may be generated by context alone, raising the possibility of misattribution to pharmacology and limiting the internal validity of psychedelic research. Moreover, experiences such as ego dissolution and altered states of consciousness, which are often associated with treatment outcomes (Romeo et al., 2025), may not only be prognostic of clinical response but may also represent modifiable targets whose emergence, intensity, and duration can be shaped by contextual factors (Stein & Terhune, 2025), thereby offering a potential pathway for enhancing therapeutic outcomes.

### Conclusions

The present experiment demonstrates that a psychedelic context alone can elicit both psychedelic effects and side effects in the absence of pharmacological action. Individual difference factors previously linked to psychedelic outcomes accounted for a modest proportion of variance but consistently predicted placebo-psychedelic effects. These findings highlight the need to shift away from viewing context solely as a moderator of psychedelic experiences to recognising it as a causal influence on such outcomes. Furthermore, these results highlight a salient asymmetry: although pharmacological action cannot readily be disentangled from its context, psychedelic contexts can be dissociated from pharmacological agents and investigated independently, providing a fertile avenue for research on psychedelic experiences.

## Acknowledgements

ChatGPT (OpenAI) was used as a support tool during data analysis and manuscript preparation. The authors retain full responsibility for the integrity, interpretation, and reporting of the work.

## Disclosures

MVS, SC, QD, and DBT have no conflicts of interest to declare. MB receives royalty fees from Taylor and Francis for a psychopharmacology textbook. He has worked as a medic on psychopharmacological trials sponsored by King’s College London, Compass Pathways, Beckley PsyTech, Multidisciplinary Association for Psychedelic Studies, and Jannsen.

## Data Availability Statement

Links to the pre-registration and data are presented in the manuscript.

## Funding sources

The research was supported by the International Society for the Study of Trauma and Dissociation (MVS) and the Gyllenbergs Foundation (MVS & DBT). The funding source had no role in study design, data interpretation, or reporting. MB is a Wellcome Trust Doctoral Clinical Research Fellow (227515/Z/23/Z).

## Author contributions

MVS contributed to study design, data collection, data analyses, drafting the paper, and integration of suggested revisions from co-authors of the final version. MB contributed to study design, data collection, and approval of the final manuscript version. SC and QD contributed to study design and approval of the final manuscript. DBT contributed to and supervised all phases of the process including design, data collection, data analyses, drafting of the paper, and approval of final version.

## Notes

### Author Declarations

This experiment was approved by the King's College London Research Ethics Committee.

## References

Aday, J. S., Davis, A. K., Mitzkovitz, C. M., Bloesch, E. K., & Davoli, C. C. (2021). Predicting reactions to psychedelic drugs: A systematic review of states and traits related to acute drug effects. ACS Pharmacol Transl Sci, 4(2), 424–435.

Aday, J. S., Heifets, B. D., Pratscher, S. D., Bradley, E., Rosen, R., & Woolley, J. D. (2022). Great expectations: Recommendations for improving the methodological rigor of psychedelic clinical trials. Psychopharmacology (Berl), 239(6), 1989–2010.

Aday, J. S., Simonsson, O., Schindler, E. A. D., & D’Souza, D. C. (2025). Addressing blinding in classic psychedelic studies with innovative active placebos. International Journal of Neuropsychopharmacology, 28(4), pyaf023.

Andersen, K. A. A., Carhart-Harris, R., Nutt, D. J., & Erritzoe, D. (2021). Therapeutic effects of classic serotonergic psychedelics: A systematic review of modern-era clinical studies. Acta Psychiatr Scand, 143(2), 101–118.

Ashar, Y. K., Chang, L. J., & Wager, T. D. (2017). Brain mechanisms of the placebo effect: An affective appraisal account. Annual Review of Clinical Psychology, 13(1), 73–98.

Bălăeţ, M. (2024). Considering the nocebo effect in the psychedelic discourse. Journal of Psychedelic Studies, 8(2), 286–291.

Barsky, A. J., Wyshak, G., & Klerman, G. L. (1990). The somatosensory amplification scale and its relationship to hypochondriasis. J Psychiatr Res, 24(4), 323–334.

Benedetti, F., & Dogue, S. (2015). Different placebos, different mechanisms, different outcomes: Lessons for clinical trials. PLoS One, 10(11), e0140967.

Benjamini, Y., & Yekutieli, D. (2001). The control of the false discovery rate in multiple testing under dependency. The Annals of Statistics, 29(4), 1165–1188, 1124.

Bhatt, K. V., Asuncion, J. D., Alam, A., Zisook, S., & Stahl, S. M. (2025). Should we skip the trip? Clinical implications of psychedelic-associated subjective effects and the potential role of non-hallucinogenic alternatives. General Hospital Psychiatry, 96, 116–120.

Bieniek, H., & Bąbel, P. (2023). Placebo hypoalgesia induced by operant conditioning: A comparative study on the effects of verbal, token-based, and social rewards and punishers. Scientific Reports, 13(1), 20346.

Blythe, J. S., Thomaidou, M. A., Peerdeman, K. J., van Laarhoven, A. I. M., van Schothorst, M. M. E., Veldhuijzen, D. S., & Evers, A. W. M. (2023). Placebo effects on cutaneous pain and itch: A systematic review and meta-analysis of experimental results and methodology. Pain, 164(6), 1181–1199.

Brake, B., Wieder, L., Hughes, N., Lalinde, I. S., Marr, D., Geagea, D., Pick, S., Reinders, A., Kamboj, S. K., Thompson, T., & Terhune, D. B. (2025). The induction of dissociative states: A meta-analysis. Biol Psychiatry Glob Open Sci, 5(4), 100521.

Bräscher, A. K., Sütterlin, S., Scheuren, R., Van den Bergh, O., & Witthöft, M. (2020). Somatic symptom perception from a predictive processing perspective: An empirical test using the thermal grill illusion. Psychosom Med, 82(7), 708–714.

Breeksema, J. J., Kuin, B. W., Kamphuis, J., van den Brink, W., Vermetten, E., & Schoevers, R. A. (2022). Adverse events in clinical treatments with serotonergic psychedelics and mdma: A mixed-methods systematic review. J Psychopharmacol, 36(10), 1100–1117.

Bremner, J. D., Krystal, J. H., Putnam, F. W., Southwick, S. M., Marmar, C., Charney, D. S., & Mazure, C. M. (1998). Measurement of dissociative states with the clinician-administered dissociative states scale (cadss). J Trauma Stress, 11(1), 125–136.

Browne, K., Bałkowiec-Iskra, E., Elferink, A., Haberkamp, M., Straus, S., Sepodes, B., Silva, F., Butlen-Ducuing, F., Jansen, J., Silva, I., Balabanov, P., Burgos, J. G., & Thirstrup, S. (2025). Applying the eu regulatory framework to determine the benefit-risk profile of psychedelics. ACS Pharmacol Transl Sci, 8(8), 2830–2838.

Bschor, T., Nagel, L., Unger, J., Schwarzer, G., & Baethge, C. (2024). Differential outcomes of placebo treatment across 9 psychiatric disorders: A systematic review and meta-analysis. JAMA Psychiatry, 81(8), 757–768.

Burke, M. J., & Blumberger, D. M. (2021). Caution at psychiatry’s psychedelic frontier. Nat Med, 27(10), 1687–1688.

Burke, M. J., Sandra, D. A., Peciña, M., Olson, J. A., Mollica, A., Butler, M., Moss, J. H., Nicholson, T. R., Wager, T. D., & Kaptchuk, T. J. (2026). Harnessing placebo effects and mitigating nocebo effects: Implications for clinical practice in psychiatry and medicine. Lancet Psychiatry, 13(5), 413–425.

Butler, M., Jelen, L., & Rucker, J. (2022). Expectancy in placebo-controlled trials of psychedelics: If so, so what? Psychopharmacology (Berl), 239(10), 3047–3055.

Carhart-Harris, R. L., & Goodwin, G. M. (2017). The therapeutic potential of psychedelic drugs: Past, present, and future. Neuropsychopharmacology, 42(11), 2105–2113.

Carhart-Harris, R. L., Roseman, L., Haijen, E., Erritzoe, D., Watts, R., Branchi, I., & Kaelen, M. (2018). Psychedelics and the essential importance of context. Journal of Psychopharmacology, 32(7), 725–731.

Carlson, E. B., & Putnam, F. W. (1993). An update on the dissociative experiences scale. Dissociation: Progress in the Dissociative Disorders, 6(1), 16–27.

Colloca, L. (2024). The nocebo effect. Annu Rev Pharmacol Toxicol, 64, 171–190.

Colloca, L., & Barsky, A. J. (2020). Placebo and nocebo effects. New England Journal of Medicine, 382(6), 554–561.

Colloca, L., & Fava, M. (2024). What should constitute a control condition in psychedelic drug trials? Nature Mental Health, 2(10), 1152–1160.

Colloca, L., & Miller, F. G. (2011). How placebo responses are formed: A learning perspective. Philos Trans R Soc Lond B Biol Sci, 366(1572), 1859–1869.

Condon, L. P., & Lynn, S. J. (2014). State and trait dissociation: Evaluating convergent and discriminant validity. Imagination, Cognition and Personality, 34(1), 25–37.

Corlett, C. E., Alldredge, C. T., & Elkins, G. R. (2025). Feasibility of a hypnosis intervention for a mystical experience. International Journal of Clinical and Experimental Hypnosis, 1-18.

Corneille, O., & Gawronski, B. (2024). Self-reports are better measurement instruments than implicit measures. Nature Reviews Psychology, 3(12), 835–846.

Corsi, N., & Colloca, L. (2017). Placebo and nocebo effects: The advantage of measuring expectations and psychological factors. Front Psychol, 8, 308.

Dai, R., Larkin, T. E., Huang, Z., Tarnal, V., Picton, P., Vlisides, P. E., Janke, E., McKinney, A., Hudetz, A. G., Harris, R. E., & Mashour, G. A. (2023). Classical and non-classical psychedelic drugs induce common network changes in human cortex. NeuroImage, 273, 120097.

Das, R. K., Tamman, A., Nikolova, V., Freeman, T. P., Bisby, J. A., Lazzarino, A. I., & Kamboj, S. K. (2016). Nitrous oxide speeds the reduction of distressing intrusive memories in an experimental model of psychological trauma. Psychol Med, 46(8), 1749–1759.

De Pascalis, V., Chiaradia, C., & Carotenuto, E. (2002). The contribution of suggestibility and expectation to placebo analgesia phenomenon in an experimental setting. Pain, 96(3), 393–402.

Deeley, Q., Oakley, D. A., Toone, B., Giampietro, V., Brammer, M. J., Williams, S. C., & Halligan, P. W. (2012). Modulating the default mode network using hypnosis. Int J Clin Exp Hypn, 60(2), 206–228.

Deeley, Q., Walsh, E., Oakley, D. A., Bell, V., Koppel, C., Mehta, M. A., & Halligan, P. W. (2013). Using hypnotic suggestion to model loss of control and awareness of movements: An exploratory fmri study. PLoS One, 8(10), e78324.

Dittrich, A. (1998). The standardized psychometric assessment of altered states of consciousness (ascs) in humans. Pharmacopsychiatry, 31 *Suppl 2*, 80–84.

Enck, P., Bingel, U., Schedlowski, M., & Rief, W. (2013). The placebo response in medicine: Minimize, maximize or personalize? Nat Rev Drug Discov, 12(3), 191–204.

Erritzoe, D., Barba, T., Benway, T., Joel, Z., Good, M., Layzell, M., Jones, M. B., Campbell, G., Murphy-Beiner, A., Rands, P., Boyce, M., Topping, H., Weiss, B., Timmermann, C., Nutt, D., Carhart-Harris, R., Routledge, C., & James, E. (2026). A short-acting psychedelic intervention for major depressive disorder: A phase iia randomized placebo-controlled trial. Nature Medicine, 32(2), 591–598.

Evans, J., & Lynn, S. J. (2021). Effects of hypnosis, suggestion, and implementation intention instructions on mystical-type experiences: A replication and extension. Psychology of Consciousness: Theory, Research, and Practice.

Evans, J., Robinson, O. C., Argyri, E. K., Suseelan, S., Murphy-Beiner, A., McAlpine, R., Luke, D., Michelle, K., & Prideaux, E. (2023). Extended difficulties following the use of psychedelic drugs: A mixed methods study. PLoS One, 18(10), e0293349.

Felitti, V. J., Anda, R. F., Nordenberg, D., Williamson, D. F., Spitz, A. M., Edwards, V., Koss, M. P., & Marks, J. S. (1998). Relationship of childhood abuse and household dysfunction to many of the leading causes of death in adults. The adverse childhood experiences (ace) study. Am J Prev Med, 14(4), 245–258.

Friedman, J. H., Hastie, T., & Tibshirani, R. (2010). Regularization paths for generalized linear models via coordinate descent. Journal of Statistical Software, 33(1), 1–22.

Glennon, M. J., Bird, C. I., Yadav, P., Kleine, P., Suseelan, S., Boman-Markaki, C., Kotoula, V., Butler, M., Leech, R., & Roseman, L. (2025). How to set up a psychedelic study: Unique considerations for research involving human participants. arXiv preprint arXiv:2503.22808.

Glicksohn, J., & Barrett, T. R. (2003). Absorption and hallucinatory experience. Applied Cognitive Psychology, 17(7), 833–849.

Golden, T. L., Magsamen, S., Sandu, C. C., Lin, S., Roebuck, G. M., Shi, K. M., & Barrett, F. S. (2022). Effects of setting on psychedelic experiences, therapies, and outcomes: A rapid scoping review of the literature. In F. S. Barrett & K. H. Preller (Eds.), Disruptive psychopharmacology (pp. 35–70). Springer International Publishing. 10.1007/7854_2021_298

Granqvist, P., Fredrikson, M., Unge, P., Hagenfeldt, A., Valind, S., Larhammar, D., & Larsson, M. (2005). Sensed presence and mystical experiences are predicted by suggestibility, not by the application of transcranial weak complex magnetic fields. Neurosci Lett, 379(1), 1–6.

Gudjonsson, G. H. (1989). Compliance in an interrogative situation: A new scale. Personality and Individual Differences, 10(5), 535–540.

Hartogsohn, I. (2016). Set and setting, psychedelics and the placebo response: An extra-pharmacological perspective on psychopharmacology. Journal of Psychopharmacology, 30(12), 1259–1267.

Hedges, L. V. (1981). Distribution theory for glass’s estimator of effect size and related estimators. Journal of Educational Statistics, 6(2), 107–128.

Hovmand, O. R., Poulsen, E. D., & Arnfred, S. (2024). Assessment of the acute subjective psychedelic experience: A review of patient-reported outcome measures in clinical research on classical psychedelics. Journal of Psychopharmacology, 38(1), 19–32.

Hovmand, O. R., Poulsen, E. D., Arnfred, S., & Storebø, O. J. (2023). Risk of bias in randomized clinical trials on psychedelic medicine: A systematic review. Journal of Psychopharmacology, 37(7), 649–659.

Hróbjartsson, A., & Gøtzsche, P. C. (2010). Placebo interventions for all clinical conditions. Cochrane Database Syst Rev, 2010(1), Cd003974.

Jamieson, G. A. (2005). The modified tellegen absorption scale: A clearer window on the structure and meaning of absorption. Australian Journal of Clinical and Experimental Hypnosis, 33(2), 119.

Kamboj, S. K., Gong, A. T., Sim, Z., Rashid, A. A., Baba, A., Iskandar, G., Das, R. K., & Curran, H. V. (2020). Reduction in the occurrence of distressing involuntary memories following propranolol or hydrocortisone in healthy women. Psychol Med, 50(7), 1148–1155.

Kaptchuk, T. J., Goldman, P., Stone, D. A., & Stason, W. B. (2000). Do medical devices have enhanced placebo effects? J Clin Epidemiol, 53(8), 786–792.

Kaptchuk, T. J., Kelley, J. M., Conboy, L. A., Davis, R. B., Kerr, C. E., Jacobson, E. E., Kirsch, I., Schyner, R. N., Nam, B. H., Nguyen, L. T., Park, M., Rivers, A. L., McManus, C., Kokkotou, E., Drossman, D. A., Goldman, P., & Lembo, A. J. (2008). Components of placebo effect: Randomised controlled trial in patients with irritable bowel syndrome. Bmj, 336(7651), 999–1003.

Kronenberg, G., Schoretsanitis, G., Seifritz, E., & Olbrich, S. (2024). The boon and bane of nitrous oxide. European Archives of Psychiatry and Clinical Neuroscience.

Kunkel, A., Schmidt, K., Hartmann, H., Strietzel, T., Sperzel, J.-L., Wiech, K., & Bingel, U. (2025). Nocebo effects are stronger and more persistent than placebo effects in healthy individuals. In: eLife Sciences Publications, Ltd.

Leary, T., Litwin, G. H., & Metzner, R. (1963). Reaction to psilocybin administered in a supportive environment. The Journal of Nervous and Mental Disease, 137(6).

Lii, T. R., Smith, A. E., Flohr, J. R., Okada, R. L., Nyongesa, C. A., Cianfichi, L. J., Hack, L. M., Schatzberg, A. F., & Heifets, B. D. (2023). Randomized trial of ketamine masked by surgical anesthesia in patients with depression. Nature Mental Health, 1(11), 876–886.

Little, R. J. A. (1988). A test of missing completely at random for multivariate data with missing values. Journal of the American Statistical Association, 83(404), 1198–1202.

Luoma, J. B., Chwyl, C., Bathje, G. J., Davis, A. K., & Lancelotta, R. (2020). A meta-analysis of placebo-controlled trials of psychedelic-assisted therapy. J Psychoactive Drugs, 52(4), 289–299.

Lüönd, A. M., Ayas, G., Bachem, R., Carranza-Neira, J., Eberle, D. J., Fares-Otero, N. E., Hashim, M., Iqbal, N., Jenkins, D., Kamari Songhorabadi, S., Ledermann, K., Makhashvili, N., Martin-Soelch, C., Nebioğlu, E., Oe, M., Olayinka, J. N., Olff, M., Picot, L., Seedat, S., . . . Ceylan, D. (2025). Childhood maltreatment and somatic symptoms in adulthood: Establishing a new research pathway. Neuropsychobiology, 84(2), 113–120.

Lynn, S. J., & Evans, J. (2017). Hypnotic suggestion produces mystical-type experiences in the laboratory: A demonstration proof. Psychology of Consciousness: Theory, Research, and Practice, 4(1), 23–37.

Mattes, A., & Roheger, M. (2020). Nothing wrong about change: The adequate choice of the dependent variable and design in prediction of cognitive training success. BMC Medical Research Methodology, 20(1), 296.

Meeuwis, S. H., Wasylewski, M. T., Bajcar, E. A., Bieniek, H., Adamczyk, W. M., Honcharova, S., Di Nardo, M., Mazzoni, G., & Babel, P. (2023). Learning pain from others: A systematic review and meta-analysis of studies on placebo hypoalgesia and nocebo hyperalgesia induced by observational learning. Pain.

Mehmood, M. K., Bremler, R., Spriggs, M. J., Kettner, H., Roseman, L., Damgaard, L., Carhart-Harris, R., Erritzoe, D., & Zeifman, R. J. (2025). Ceremonial psychedelic experiences and changes in mental health outcomes in those with adverse childhood experiences. Psychedelic Medicine.

Meissner, K., Fässler, M., Rücker, G., Kleijnen, J., Hróbjartsson, A., Schneider, A., Antes, G., & Linde, K. (2013). Differential effectiveness of placebo treatments: A systematic review of migraine prophylaxis. JAMA Intern Med, 173(21), 1941–1951.

Mestre, T. (2020). Chapter six - nocebo and lessebo effects. In N. P. Witek, C. G. Goetz, & G. T. Stebbins (Eds.), International review of neurobiology (Vol. 153, pp. 121–146). Academic Press. 10.1016/bs.irn.2020.04.005

Nour, M. M., Evans, L., Nutt, D., & Carhart-Harris, R. L. (2016). Ego-dissolution and psychedelics: Validation of the ego-dissolution inventory (edi). Front Hum Neurosci, 10, 269.

Olofsson, M., Osika, W., Goldberg, S. B., Hendricks, P. S., Petrovic, P., White, T., Stenfors, C. U. D., Chaturvedi, S., & Simonsson, O. (2026). Difficulties following naturalistic psychedelic use and associations with adverse childhood experiences. International Journal of Drug Policy, 148, 105105.

Olson, J. A., Suissa-Rocheleau, L., Lifshitz, M., Raz, A., & Veissière, S. P. L. (2020). Tripping on nothing: Placebo psychedelics and contextual factors. Psychopharmacology (Berl), 237(5), 1371–1382.

Owens, J. E., & Menard, M. (2011). The quantification of placebo effects within a general model of health care outcomes. J Altern Complement Med, 17(9), 817–821.

Parsons, R. D., Bergmann, S., Wiech, K., & Terhune, D. B. (2021). Direct verbal suggestibility as a predictor of placebo hypoalgesia responsiveness. Psychosom Med, 83(9), 1041–1049.

Peerdeman, K. J., van Laarhoven, A. I. M., Keij, S. M., Vase, L., Rovers, M. M., Peters, M. L., & Evers, A. W. M. (2016). Relieving patients’ pain with expectation interventions: A meta-analysis. Pain, 157(6), 1179–1191.

Piazza, G. G., Iskandar, G., Hennessy, V., Zhao, H., Walsh, K., McDonnell, J., Terhune, D. B., Das, R. K., & Kamboj, S. K. (2022). Pharmacological modelling of dissociation and psychosis: An evaluation of the clinician administered dissociative states scale and psychotomimetic states inventory during nitrous oxide (’laughing gas’)-induced anomalous states. Psychopharmacology (Berl), 239(7), 2317–2329.

Pronovost-Morgan, C., Greenway, K. T., & Roseman, L. (2025). An international delphi consensus for reporting of setting in psychedelic clinical trials. Nat Med, 31(7), 2186–2195.

Pronovost-Morgan, C., Hartogsohn, I., & Ramaekers, J. G. (2023). Harnessing placebo: Lessons from psychedelic science. J Psychopharmacol, 37(9), 866–875.

Prugger, J., Derdiyok, E., Dinkelacker, J., Costines, C., & Schmidt, T. T. (2022). The altered states database: Psychometric data from a systematic literature review. Scientific Data, 9(1), 720.

R Core Team. (2023). R: A language and environment for statistical computing (Version 4.3.1). R Foundation for Statistical Computing. https://www.R-project.org/

Rief, W., Barsky, A. J., Glombiewski, J. A., Nestoriuc, Y., Glaesmer, H., & Braehler, E. (2011). Assessing general side effects in clinical trials: Reference data from the general population. Pharmacoepidemiol Drug Saf, 20(4), 405–415.

Romeo, B., Kervadec, E., Fauvel, B., Strika-Bruneau, L., Amirouche, A., Bezo, A., Piolino, P., & Benyamina, A. (2025). The intensity of the psychedelic experience is reliably associated with clinical improvements: A systematic review and meta-analysis. Neuroscience & Biobehavioral Reviews, 172, 106086.

Rooney, T., Sharpe, L., Todd, J., Tang, B., & Colagiuri, B. (2023). The nocebo effect across health outcomes: A systematic review and meta-analysis. Health Psychol.

Rooney, T., Sharpe, L., Todd, J., Tang, B., & Colagiuri, B. (2024). The nocebo effect across health outcomes: A systematic review and meta-analysis. Health Psychol, 43(1), 41–57.

Rubanets, D., Łaska, I., Kłosowska, J., Bąbel, P., & Bajcar, E. A. (2026). Verbal modeling, counterconditioning, and operant conditioning are effective in nocebo hyperalgesia attenuation. PAIN.

Rucker, J. J. (2024). Evidence versus expectancy: The development of psilocybin therapy. BJPsych Bulletin, 48(2), 110–117.

Schenk, L. A., Fadai, T., & Büchel, C. (2024). How side effects can improve treatment efficacy: A randomized trial. Brain, 147(8), 2643–2651.

Stein, M. V., Heller, M., Chapman, S., Rubin, G. J., & Terhune, D. B. (2025a). Trait responsiveness to verbal suggestions predicts nocebo responding: A meta-analysis. Br J Health Psychol, 30(1), e12774.

Stein, M. V., Heller, M., Hughes, N., Marr, D., Brake, B., Chapman, S., Rubin, G. J., & Terhune, D. B. (2025b). Moderators of nocebo effects in controlled experiments: A multi-level meta-analysis. Neuroscience & Biobehavioral Reviews, 172, 106042.

Stein, M. V., Helstrip, J., Bąbel, P., Barnier, A. J., Bingel, U., Cardeña, E., Colagiuri, B., Colloca, L., Deeley, Q., De Pascalis, V., Eisen, M. L., Faasse, K., Gabbert, F., Geer, A. L., Gudjonsson, G. H., Huneke, N. T. M., Jamieson, G. A., Jensen, K. B., Jensen, M. P., . . . Terhune, D. B. (2025, April). Developing an expert consensus definition of suggestion: A modified-delphi study [registration].

Stein, M. V., Holt, R., Wieder, L., & Terhune, D. B. (2023). Responsiveness to direct verbal suggestions and dissociation independently predict symptoms associated with environmental factors. Psychopathology, 56(4), 324–328.

Stein, M. V., & Terhune, D. B. (2025). Suggestion effects in psychedelics: Confounds and opportunities. International Journal of Clinical and Experimental Hypnosis, 1-20.

Studerus, E., Gamma, A., Kometer, M., & Vollenweider, F. X. (2012). Prediction of psilocybin response in healthy volunteers. PLoS One, 7(2), e30800.

Szigeti, B., & Heifets, B. D. (2024). Expectancy effects in psychedelic trials. Biol Psychiatry Cogn Neurosci Neuroimaging, 9(5), 512–521.

Szigeti, B., Nutt, D., Carhart-Harris, R., & Erritzoe, D. (2023). The difference between ‘placebo group’ and ‘placebo control’: A case study in psychedelic microdosing. Scientific Reports, 13(1), 12107.

Szigeti, B., Weiss, B., Rosas, F. E., Erritzoe, D., Nutt, D., & Carhart-Harris, R. (2024). Assessing expectancy and suggestibility in a trial of escitalopram v. Psilocybin for depression. Psychol Med, 1–8.

Taillefer de Laportalière, T., Jullien, A., Yrondi, A., Cestac, P., & Montastruc, F. (2023). Reporting of harms in clinical trials of esketamine in depression: A systematic review. Psychol Med, 53(10), 4305–4315.

Tellegen, A., & Atkinson, G. (1974). Openness to absorbing and self-altering experiences (“absorption”), a trait related to hypnotic susceptibility. J Abnorm Psychol, 83(3), 268–277.

Terhune, D. B., & Jamieson, G. A. (2021). Hallucinations and the meaning and structure of absorption. Proceedings of the National Academy of Sciences, 118(32), e2108467118.

Terhune, D. B., & Smith, M. D. (2006). The induction of anomalous experiences in a mirror-gazing facility: Suggestion, cognitive perceptual personality traits and phenomenological state effects. Journal of Nervous and Mental Disease, 194(6), 415–421.

Timmermann, C., Bauer, P. R., Gosseries, O., Vanhaudenhuyse, A., Vollenweider, F., Laureys, S., Singer, T., Antonova, E., & Lutz, A. (2023). A neurophenomenological approach to non-ordinary states of consciousness: Hypnosis, meditation, and psychedelics. Trends in Cognitive Sciences, 27(2), 139–159.

van Buuren, S., & Groothuis-Oudshoorn, K. (2011). Mice: Multivariate imputation by chained equations in r. Journal of Statistical Software, 45(3), 1–67.

van Elk, M. (2015). An eeg study on the effects of induced spiritual experiences on somatosensory processing and sensory suppression. Journal for the Cognitive Science of Religion, 2, 121–157.

van Elk, M., & Fried, E. I. (2023). History repeating: Guidelines to address common problems in psychedelic science. Therapeutic Advances in Psychopharmacology, 13, 20451253231198466.

Viljoen, G., Walter, H., Bendau, A., Koslowski, M., & Betzler, F. (2025). Predictors of therapeutic response to psychedelic-assisted therapy: A systematic review. Journal of Psychopharmacology, 02698811251389581.

Vizeli, P., Studerus, E., Holze, F., Schmid, Y., Dolder, P. C., Ley, L., Straumann, I., Becker, A. M., Müller, F., Arikci, D., & Liechti, M. E. (2024). Pharmacological and non-pharmacological predictors of the lsd experience in healthy participants. Translational Psychiatry, 14(1), 357.

Wager, T. D., & Atlas, L. Y. (2015). The neuroscience of placebo effects: Connecting context, learning and health. Nat Rev Neurosci, 16(7), 403–418.

Wen, A., Singhal, N., Jones, B. D., Zeifman, R. J., Mehta, S., Shenasa, M. A., Blumberger, D. M., Daskalakis, Z. J., & Weissman, C. R. (2024). A systematic review of study design and placebo controls in psychedelic research. Psychedelic Medicine, 2(1), 15–24.

Weng, L., Peerdeman, K. J., van Laarhoven, A. I. M., & Evers, A. W. M. (2025). Generalisation of placebo and nocebo effects: Current knowledge and future directions. European Journal of Pain, 29(6), e70018.

Wiech, K., Hartmann, H., & Bingel, U. (2024). Side-effects are often a curse. Can they also be a blessing? Brain, 147(8), 2598–2600.

Wieder, L., & Terhune, D. B. (2019). Trauma and anxious attachment influence the relationship between suggestibility and dissociation: A moderated-moderation analysis. Cognitive Neuropsychiatry, 24(3), 191–207.

Winkler, J., Kanouse, D., & Ware, J. (1982). Controlling for acquiescence response set in scale development. Journal of Applied Psychology, 67, 555–561.

Wittmann, M., Carter, O., Hasler, F., Cahn, B. R., Grimberg, U., Spring, P., Hell, D., Flohr, H., & Vollenweider, F. X. (2007). Effects of psilocybin on time perception and temporal control of behaviour in humans. J Psychopharmacol, 21(1), 50–64.

Wongpaiboon, M. P., Shelton, J. L., Rendon, E. D., & Vail, M. (2025). A systematic review of the efficacy, safety, and tolerability of psychedelics in the treatment of post-traumatic stress disorder. Current Treatment Options in Psychiatry, 13(1), 2.

Yaden, D. B., & Griffiths, R. R. (2021). The subjective effects of psychedelics are necessary for their enduring therapeutic effects. ACS Pharmacol Transl Sci, 4(2), 568–572.

Yanakieva, S., Polychroni, N., Family, N., Williams, L. T. J., Luke, D. P., & Terhune, D. B. (2019). The effects of microdose lsd on time perception: A randomised, double-blind, placebo-controlled trial. Psychopharmacology (Berl), 236(4), 1159–1170.

Zorumski, C. F., Cichon, J., Izumi, Y., Zeffiro, T., Mennerick, S., Nagele, P., & Conway, C. R. (2025). Rapid antidepressant potential of nitrous oxide: Current state and major questions. Molecular Psychiatry.

Zou, H., & Hastie, T. (2005). Regularization and variable selection via the elastic net. Journal of the Royal Statistical Society Series B: Statistical Methodology, 67(2), 301–320.

