## Supplementary materials for "Tripping on context: Characteristics and predictors of placebo and nocebo psychedelic effects"

**For the purposes of open access, the author has applied a Creative Commons Attribution (CC BY) license to any Accepted Author manuscript version arising from this submission.**

### Supplementary methods

*Participants*

The sample size was determined *a priori* (80% power, α = .05) to detect a moderate placebo/nocebo effect in the primary analyses and to detect a moderate effect size ($f^2 = .20$; corresponding to ~15% of the variance) in a multiple regression analysis with six predictors. Based on this analysis, 75 participants were required. To account for attrition, we recruited 80 participants.

One hundred and sixty-seven people were screened via telephone, of whom 118 were deemed eligible to participate. Among these, 38 did not take part in the experiment, resulting in a total sample size of 80 participants. Two participants were excluded from the data analyses; one did not complete the second inhalation period and the other was excluded due to English language comprehension.

Of the 78 participants, 27 (35%) identified as White (including British, Irish, European, American, South African, and Greek), 20 (26%) as Asian/Asian British (South Asian; Indian, Pakistani, Bangladeshi, Tamil), 11 (14%) as Asian/Asian British (Other Asian; e.g., Burmese, unspecified Asian), eight (10%) as Asian/Asian British (Chinese), 6 (8%) as Mixed/Multiple ethnic groups, three (4%) as Black/African/Caribbean/Black British, and two (3%) did not to disclose their ethnicity.

Age, post-secondary education data, and previous nitrous oxide ($N_2O$) use data were available for 77 participants due to one participant who elected not to respond.

### Measures

*Trait measures*

The *Adverse Childhood Experience Checklist* (ACE) (Felitti et al., 1998) is a 10-item self-report scale that indexes exposure to potentially traumatic events in childhood, including physical, emotional, sexual abuse, as well as physical and emotional maltreatment. Participants rate each item in a binary fashion (0 = no, 1 = yes) with total scores ranging from 0 to 10. All age of event questions were omitted. The ACE demonstrates reliable psychometric properties for retrospective assessment of childhood abuse and maltreatment (Felitti et al., 1998) and displayed marginal internal consistency in the present sample (Cronbach's α = .68).

The *Acquiescence Response Set* (ARS) (Winkler et al., 1982) assesses respondents' tendency to agree with questionnaire statements regardless of content. Each item is rated as a logical opposite in a binary fashion (e.g., 0 = disagree, 1 = agree). The ARS has historically demonstrated poor psychometrics properties (Winkler et al., 1982), which were observed in our sample (α = .17). Owing to the poor internal consistency, we deviated from our pre-registration (see main paper) and omitted this measure from all analyses.

The *Brief Suggestibility Scale* (BSS) (Wieder & Terhune, 2019) is a behavioural measure used to measure non-hypnotic responsiveness to direct verbal suggestions. Participants listened via headphones to six verbal suggestions that were drawn from standardized hypnotic suggestibility scales (arm heaviness, dream, hands moving together, eye catalepsy, arm paralysis, and music hallucination) (Bowers, 1998; Shor, 1962; Weitzenhoffer, 1962), which were followed by brief behavioural tests. Participants then rated their responsiveness to each suggestion based on their behavioural response using a continuous visual analogue scale (0 = no response to 1 = complete response) with the aid of item-specific behavioural response anchors and their experience of involuntariness (0 = did not experience at all, 1 = voluntary and 5 = involuntary; (Bowers, 1981), in order to capture the classic suggestion effect and correct for compliance (Bowers et al., 1988). Corrections for compliance were made by computing the average of z-transformed behavioural and involuntariness scores (BSS-C). All measures demonstrated marginal-to-acceptable internal consistency in the present sample (αs: behavioural = .68; involuntariness = .70).

The *Dissociative Experiences Scale-II* (DES) (Carlson & Putnam, 1993) is a 28-item self-report measure of dissociative tendencies. Using an 11-point-scale, participants are asked to rate the frequency with which they have had different dissociative experiences (0% = never to 100% = always). Along with total scores, the DES yields three subscale scores: dissociative absorption (nine items), amnesia (eight items), and depersonalization/derealization (six items). The DES has demonstrated strong test-retest reliability and construct-validity (Carlson & Putnam, 1993) and had strong internal consistency in the present sample (αs: DES = .91; absorption = .84; amnesia = .68; depersonalization-derealization = .85).

The *Somatosensory Amplification Scale* (SSAS) (Barsky et al., 1990) assesses the respondents' tendency to view and experience benign somatic and visceral sensations as unusually intense and alarming. Using a five-point Likert, respondents indicate the extent to which they agree with a given statement (e.g., 'I hate to be too hot or too cold'). The SSAS is often used in nocebo and nocebo-germane research (Doering et al., 2016; Skovbjerg et al., 2010), and demonstrated marginal internal consistency in the present sample (α = .67).

The *Gudjonsson Compliance Scale* (GCS) (Gudjonsson, 1989) is a 20-item assessment of respondents' tendency to comply with others. Participants rate a given statement (e.g., "I tend to give in to people who insist that they are right") in a binary fashion (0 = false, 1 = true) with total scores ranging from 0 to 20. It has validated psychometric properties (Gudjonsson, 1989) and has demonstrated good internal consistency (Nash et al., 2022), which was replicated in our sample (α = .70).

*State measures*

The *5-Dimensional Altered States of Consciousness Questionnaire* (5D-ASC) (Dittrich, 1998) is a 94-item measure assessing alterations in consciousness, such as feelings of oneness, positive or negative shifts in affect, and sensory distortions. The items are answered on a visual analogue scale ranging from 0 to 100 (0 = 'not more than usual', 100 = 'much more than usual'). The 5D-ASC comprises five subscales assessing classic psychedelic experiences: oceanic boundlessness (OBN), dread of ego dissolution (DED), visionary restructuralization (VRS), auditory alterations (AUA), and vigilance reduction (VIR). It is commonly used in classic (i.e., serotonergic) and non-classic (e.g., ketamine) psychedelic research (Prugger et al., 2022) and has strong construct validity and reliability (Studerus et al., 2010). The total scale and subscales yielded good psychometric properties in our sample (αs: placebo: total=.97, OBN=.96, DED=.96, VRS=.83, AUA=.87, VIR=.91; control: total=.96, OBN=.85, DED=.95, VRS=.70, AUA=.79, VIR=.91).

The *Clinician Administered Dissociative States Scale* (CADSS) (Bremner et al., 1998) is a 19-item self-report assessment of dissociation at the time of the assessment (i.e., state dissociation). Using a five-point scale, participants are asked to rate their experience of dissociation (0 = 'not at all' to 4 = 'extremely'). In addition to a total score, the CADSS includes three subscales indexing dissociative amnesia, depersonalization, and derealisation. The CADSS has been widely used in experimental (Morgan et al., 2004) and clinical studies of ketamine (Niciu et al., 2018) and nitrous oxide (Brake et al., 2025; Piazza et al., 2022). It has strong psychometric properties (Piazza et al., 2022) and all scales demonstrated good internal consistency in our sample except the amnesia subscale (αs: placebo: total=.93, amnesia=.59, depersonalization=.91, derealization=.87; control: total=.89, amnesia=.57, depersonalization=.72, derealization=.85).

The *Ego-Dissolution Inventory* (EDI) (Nour et al., 2016) is an eight-item measure assessing respondents' perceived self-boundaries in response to psychoactive substances. The items are answered on a visual analogue scale (0 = 'no, not more than usual, 1 = 'yes, entirely or completely'). The EDI is commonly used in classic (Jensen et al., 2022) and non-classic psychedelic studies (Earleywine et al., 2022), and has good construct validity and reliability (Nour et al., 2016). In our sample, the EDI demonstrated good internal consistency in both conditions (αs: placebo = .93; control = .70).

The *General Assessment of Side Effects* (GASE) (Rief et al., 2011) consists of 36-items that ask about physical and emotional symptoms during the last week. Using a four-point Likert, respondents rate the symptom severity (0 = not present, 3 = severe). We modified the temporal reference point to the preceding inhalation phase and omitted one item about suicidality. GASE scores reflected summed symptom severity transformed into percentage scores to account for sex-specific item differences. The GASE is commonly used to measure side effects in nocebo experiments and related studies however, its frequent modifications can complicate comparisons between different studies (Barnes et al., 2019). In our study, it demonstrated good internal consistency (αs: placebo = .85; control = .84).

*Manipulation checks*

The *Guess of Treatment Questionnaire* (GoTQ) (Szigeti et al., 2023) was used to assess participants' blinding to the experimental condition. The GoTQ consists of five items (all rated on a VAS from 0% to 100%) and asks participants to rate the likelihood they were administered an active treatment or placebo, their confidence in this rating, and to what extent their selection was based on experience of therapeutic effects or side effects.

Participants completed a single-item compliance check measured on a 5-point Likert scale. The scale ranged from 1 ('All my responses were based solely on my experiences in the experiment') to 5 ('All my responses were based solely on how I thought you expected me to respond'), with higher scores reflecting greater compliant responding.

*Screening*

To enhance the credibility of the advertised experiment's aims, we maintained the same exclusion criteria as a genuine $N_2O$ study (see Kamboj et al., 2020; Das et al., 2016). As such, all participants were required to not meet the following exclusion criteria (based on self-report): a) 18 > age > 45; b) BMI > 40; c) proneness to nausea; d) any current or historical psychiatric, neurological, or medical diagnosis (inclusive of asthma and allergies); e) history of adverse reaction to $N_2O$ or any other medication; f) $N_2O$ use >4 times a week; and/or g) previous participation in a drug intervention study using $N_2O$. Additionally, we excluded current psychology students at King's College London, as students in this course had previously learned about placebo effects including in the context of psychedelic drug administration. During the screening process, participation received indirect verbal and textual suggestions within the information sheet, and during a telephone screening call.

Information sheet: *"When inhaling Entonox, many participants experience a variety of psychedelic effects including hallucinations, euphoria, changes in their perception of their body, and other types of anomalous experiences. However, bear in mind that these experiences are different for everyone, some people might have an intense experience while others might not. Entonox is completely safe but some participants also experience minor side effects including nausea, dizziness, fatigue, and other minor side effects."*

During phone screening: "*...[$N_2O$] can have euphoric effects when you inhale it. Often it has no side effects or, in some cases, it can have some side effects including fatigue, nausea, dizziness and other mild side effects. On rare occasions, the adverse reaction involves feeling sick and vomiting."*

*Supplementary analyses*

We made multiple deviations from our pre-registered analysis plan (https://osf.io/4ab7r). Our pre-registration specified log transformation of non-normally distributed data; however, these transformations did not meaningfully improve distributional characteristics. As the assumption of normality remained violated, we instead employed nonparametric analyses, which are more robust to such violations and less sensitive to the influence of outliers (Erceg-Hurn & Mirosevich, 2008). Among self-report measures, we identified 13 univariate outliers ($|Z| > 3$) and three multivariate outliers based on Mahalanobis distance ($p < .001$). As excluding a total of 16 participants was deemed infeasible, we deviated from our original plan and retained these outliers in the dataset. Two univariate outliers were also identified in the temporal reproduction data but were retained in the dataset to maintain consistency across analytic approaches.

Three participants were removed from relevant self-report analyses: one did not complete the ACE and GCS at baseline, one did not complete the GoTq and the compliance measure, and one provided a confidence rating of zero on the GoTq, which is equivalent to a random guess.

### Supplementary results

*Order and condition-order interactions*

Across outcomes, there were robust main effects of condition, with placebo–control differences observed for most measures (**Supplementary Table 1**). Order effects were generally small ($\eta_p^2$s < .08) and inconsistent. Significant condition × order interactions were observed for state dissociation, depersonalization and derealisation (CADSS total, CADSS Dep, CADSS Der), and ego dissolution (EDI). Decomposition of these interactions indicated that placebo–control differences were consistently larger when the placebo condition was administered prior to the control condition compared to when it was administered after the control condition.

**Supplementary Table 1**. *Results of 2 × 2 (Condition × Order) permutation ANOVAs with decomposition of significant interaction effects (N=78).*

| Outcome | Condition | | | Order | | | Interaction | | | Placebo first | | Control first | |
|---|---|---|---|---|---|---|---|---|---|---|---|---|---|
| | *F* | *p* | $\eta_p^2$ | *F* | *p* | $\eta_p^2$ | *F* | *p* | $\eta_p^2$ | *g* [95% CI] | *p* | *g* [95% CI] | *p* |
| 5D-ASC | 30.46 | <.001 | .29 | 3.59 | .058 | .05 | 4.33 | .043 | .05 | - | - | - | - |
| 5D-ASC AUA | 11.12 | <.001 | .13 | 0.65 | .44 | .01 | 0.97 | .34 | .01 | - | - | - | - |
| 5D-ASC DED | 4.31 | .038 | .05 | 0.78 | .41 | .01 | 1.33 | .26 | .02 | - | - | - | - |
| 5D-ASC OBN | 11.47 | <.001 | .13 | 5.41 | .013 | . 07 | 3.93 | .036 | .05 | - | - | - | - |
| 5D-ASC VIR | 30.68 | <.001 | .29 | 3.33 | .068 | .04 | 1.50 | .22 | .02 | - | - | - | - |
| 5D-ASC VRS | 14.80 | <.001 | .16 | 3.17 | .074 | .04 | 0.52 | .48 | .01 | - | - | - | |
| CADSS | 26.48 | <.001 | .26 | 1.68 | .21 | .02 | 11.35 | .002 | .13 | 0.77 [0.52, 1.07] | <.001 | 0.28 [-0.02, 0.60] | .08 |
| CADSS Amn | 9.74 | .003 | .11 | 0.48 | .49 | .01 | 1.89 | .17 | .02 | - | - | - | - |
| CADSS Dep | 21.10 | <.001 | .22 | 4.11 | .043 | .05 | 11.22 | <.001 | .13 | 0.69 [0.52, 0.94] | <.001 | 0.21 [-0.07, 0.55] | .21 |
| CADSS Der | 24.75 | <.001 | .25 | 1.57 | .22 | .02 | 9.58 | .003 | .11 | 0.75 [0.50, 1.07] | <.001 | 0.27 [-0.03, 0.56] | .10 |
| EDI | 12.11 | .001 | .14 | 3.11 | .079 | .04 | 10.55 | <.001 | .12 | 0.60 [0.43, 0.81] | <.001 | 0.04 [-0.30, 0.32] | .82 |
| GASE | 33.30 | <.001 | .30 | 0.26 | .62 | .00 | 4.02 | .047 | .05 | - | - | - | - |

***Notes.*** 5D-ASC = *5-Dimensional Altered States of Consciousness Rating Scale*; 5D-ASC subscales: AUA = Auditory Alterations; DED = Dread of Ego Dissolution; OBN = Oceanic Boundlessness; VIR = Vigilance Reduction; VRS = Visionary Restructuralization; CADSS = *Clinician-Administered Dissociative States Scale*; CADSS Amn = CADSS-Amnesia subscale; CADSS-Dep = CADSS Depersonalization subscale; CADSS Der = CADSS-Derealization subscale; EDI = *Ego Dissolution Inventory*; GASE = *Generic Assessment of Side Effects*. Each row represents a separate outcome variable analysed using a 2 (Condition: placebo vs. control) × 2 (Order: placebo-first vs. control-first) permutation ANOVA. Reported values are *F*-statistics, exact *p*-values, and partial eta squared ($\eta_p^2$). Interaction effects were followed up with decomposition analyses only when they remained significant after Benjamini–Hochberg FDR correction (FDR $p$ = .003). Decomposition of these interactions are shown using Hedges' *g* [95% CIs], and raw *p*-values for each order group. All effects remained significant after Benjamini–Hochberg FDR correction (FDR $p$ = .004).

*Blinding and compliance*

Participants' confidence in their condition guess was moderate in the placebo condition ($N$ = 78, $M$ = 60.38, $SD$ = 29.82, $Mdn$ = 53.5, IQR = 33.5–91), and high in the control condition ($n$ = 77, $M$ = 82.23, $SD$ = 22.68, $Mdn$ = 94.0, IQR = 60–100). As shown in **Supplementary Table 2** and **Supplementary Figure 1**, attribution ratings reflect the perceived contribution of health improvements and side effects/perceptual changes to participants' condition guesses. Thus, lower percentages in the blinded group reflect reduced attribution of guesses to the respective variable, not weaker effects.

In addition to Likert ratings, participants were provided with free-text boxes to describe factors contributing to their condition guesses. A summary of these themes is presented in **Supplementary Table 3.** Across both conditions, common themes included lack of expected changes and perceptual experiences, although blinded participants more frequently referenced information provided, whereas unblinded participants more often cited absence of expected effects. Among those who remained blinded to the placebo procedure ($n$ = 35), responses provided insight into the basis of condition guesses: approximately 29% reported relying on information provided during the procedure, whereas 23% attributed their guesses to perceptual changes.

**Supplementary Table 2**. *Blinding and unblinding descriptive statistics by condition.*

| | | Side effects and perceptual effects attribution (%) | | | Health improvement attribution (%) | | | Confidence (%)* | | |
|---|---|---|---|---|---|---|---|---|---|---|
| Awareness subgroup | *n* | *M* (*SD*) | *Med* | IQR | *M* (SD) | *Med* | IQR | *M* (*SD*) | *Med* | *IQR* |
| Placebo condition | | | | | | | | | | |
| Blinded (guessed $N_2O$) | 34 | 54.44 (37.08) | 50 | 23.25–95.25 | 15.18 (27.37) | 0 | 0–15.75 | 63.50 (29.10) | 65 | 47-91 |
| Unblinded (guessed medical air) | 43 | 68.35 (34.04) | 82 | 50–100 | 33.44 (38.75) | 12 | 0–68.50 | 58.69 (30.50) | 51 | 32.25-89.75 |
| Control condition | | | | | | | | | | |
| Incorrect (guessed $N_2O$) | 5 | 67.60 (41.22) | 81 | 59–98 | 21 (25.51) | 17 | 0–26.00 | - | - | - |
| Correct (guessed medical air) | 72 | 75.15 (32.55) | 90.5 | 50–100 | 37.83 (39.28) | 25 | 0–75.25 | - | - | - |

***Notes.*** *=Sample size differs for confidence rating descriptive statistics (blinded *n* = 35; unblinded *n* = 42). One participant did not complete the GoTQ in the control condition. Another participant was removed from the placebo condition summary due to a confidence rating of zero (equivalent to a random guess).

**Supplementary Figure 1.** *Guess of condition confidence and attribution within the placebo condition (n = 78).*

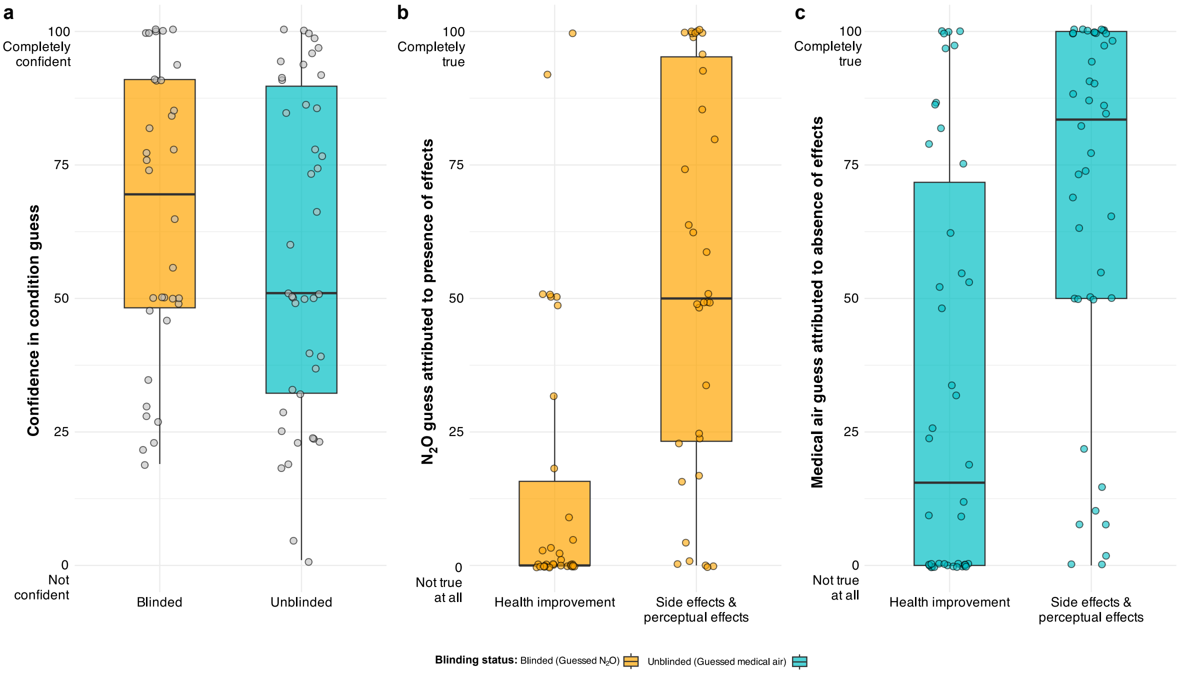


***Notes.*** Boxplots display the median and interquartile range (IQR), with individual data points overlaid. Panels represent different ratings: (a) confidence in condition guess, (b, c) guess attributed to respective presence or absence of health improvements, side effects or perceptual effects.

**Supplementary Table 3**. *Frequency of themes mentioned in free-text responses regarding the factors contributing to condition guesses presented by condition and blinding status.*

| **Condition** | ***n*** | **Reason *n* (%)** | | | | | |
|---|---|---|---|---|---|---|---|
| | | **No response** | **Context features** | **Information provided** | **Perceptual changes** | **Lack of expected changes** | **Mention placebo effect** |
| Placebo | 78 | 39 (50%) | 5 (6%) | 10 (13%) | 11 (14%) | 14 (18%) | 1 (1%) |
| Control | 78 | 44 (56%) | 6 (8%) | 11 (14%) | 2 (3%) | 17 (22%) | 0 (0%) |
| **Blinding status (placebo condition)** | | | | | | | |
| Blinded* | 35 | 12 (34%) | 4 (11%) | 10 (29%) | 8 (23%) | 4 (11%) | 1 (3%) |
| Unblinded | 43 | 27 (62%) | 1 (2%) | 0 (0%) | 3 (7%) | 10 (23%) | 0 (0%) |

***Notes.*** * = Includes participant with a confidence rating of zero (equivalent to a random guess); context features include gas exhaust hose, cylinder label, etc.; Counts exceed sample size due to participants mentioning multiple themes.

*Trait predictors of blinding status*

A series of exploratory analyses examined trait responsiveness to verbal suggestions (BSS), dissociative absorption (DES-Abs), and compliance (GCS) as predictors of blinding status using binary logistic regression (0 = unblinded, 1 = blinded). A combined model including both BSS and DES-Abs was not significant, $\chi^2(2, N = 77) = 2.25$, $p = .32$, Nagelkerke $R^2 = .04$, with neither BSS (odds ratio [OR] = 1.48 [0.82, 2.76], $p = .20$) nor DES-Abs (OR = 1.01 [0.98, 1.03], $p = .67$) significantly associated with blinding status. A final univariate model assessing GCS as a predictor indicated that higher GCS scores were not associated with increased odds of being blinded (OR = 1.05 [1.00, 1.20], $p = .45$). Taken together, these results indicate that these individual difference variables did not predict blinding status in the present sample.

*Placebo and nocebo effects as a function of condition blinding*

Condition effects stratified by blinding status are presented in **Supplementary Table 4**. Blinding integrity was considered maintained when participants in the placebo condition reported that they had received $N_2O$ and indicated a confidence rating greater than 0.

Across both blinding groups, placebo and nocebo effects (placebo-control) ranged from weak-to-large. In the blinded group, significant effects were observed for altered states of consciousness (Δ5D-ASC total) and its corresponding subscales (Δ5D-ASC AUA, DED, OBN, VIR, VRS), state dissociation (ΔCADSS total) including depersonalisation and derealisation subscales (but not amnesia), ego dissolution (ΔEDI), and side effects (ΔGASE). In the unblinded group, significant effects were observed for altered states of consciousness (Δ5D-ASC total) and three of its corresponding subscales (Δ5D-ASC OBN, VIR, VRS) and state dissociation (ΔCADSS total) and derealization, and ΔGASE, whereas effects for AUA, DED, CADSS Amnesia, CADSS Depersonalisation, and EDI were not significant. Compared to the main analyses, the magnitude of placebo and nocebo effects were somewhat attenuated in the unblinded group, with fewer outcomes reaching statistical significance and generally smaller effect sizes. However, direct comparisons between blinded and unblinded participants revealed a significant difference only for the auditory alterations subscale of the Δ5D-ASC. No other outcomes differed between groups, indicating that blinding status did not reliably moderate the magnitude of placebo and nocebo effects.

**Supplementary Table 4**. *Magnitude of condition effects (placebo – control) in self-report measures stratified by blinding status.*

| | Blinded* (*n* = 34) | | | | | Unblinded (*n* = 43) | | | | | Blinded v. unblinded (*n* = 77) | |
|---|---|---|---|---|---|---|---|---|---|---|---|---|
| Outcome | Control | Placebo | *t*(33) | *p* | *g* [95% CIs] | Control | Placebo | *t*(42) | *p* | *g* [95% CIs] | Δ*g* [95% CI] | *p* (*t-test*) |
| | *M* (*SD*) | | | | | *M* (*SD*) | | | | | | |
| 5D-ASC | 4.35 (6.88) | 9.45 (10.82) | 4.50 | <.001 | 0.55 [0.36, 0.96] | 2.40 (3.84) | 4.62 (6.11) | 2.93 | .001 | 0.43 [0.19, 0.72] | 0.12 [-0.18, 0.52] | .027 |
| 5D-ASC AUA | 1.97 (5.09) | 4.91 (10.52) | 2.77 | <.001 | 0.35 [0.24, 0.67] | 0.56 (1.57) | 1.12 (2.63) | 1.89 | .053 | 0.26 [0.03, 0.57] | 0.09 [-0.17, 0.43] | .008 |
| 5D-ASC DED | 4.18 (11.34) | 6.27 (11.99) | 2.77 | .003 | 0.18 [0.05, 0.58] | 2.15 (6.23) | 3.79 (11.03) | 1.00 | .45 | 0.18 [-0.09, 0.49] | -0.01 [-0.27, 0.48] | .92 |
| 5D-ASC OBN | 2.25 (4.68) | 8.30 (15.54) | 2.49 | <.001 | 0.52 [0.37, 0.88] | 1.32 (2.76) | 3.25 (5.80) | 2.32 | .020 | 0.42 [0.13, 0.75] | 0.10 [-0.19, 0.47] | .062 |
| 5D-ASC VIR | 16.19 (18.53) | 29.11 (21.49) | 4.34 | <.001 | 0.63 [0.36, 1.01] | 9.44 (12.84) | 16.96 (18.31) | 3.41 | .001 | 0.47 [0.23, 0.77] | 0.16 [-0.23, 0.57] | .14 |
| 5D-ASC VRS | 1.90 (3.89) | 5.84 (9.55) | 3.15 | <.001 | 0.53 [0.38, 0.86] | 1.28 (2.92) | 2.51 (5.09) | 2.08 | .041 | 0.29 [0.04, 0.58] | 0.24 [-0.05, 0.57] | .029 |
| CADSS | 4.38 (5.68) | 10.41 (10.90) | 4.47 | <.001 | 0.68 [0.44, 1.09] | 4.21 (6.01) | 6.47 (7.04) | 2.16 | .035 | 0.34 [0.05, 0.69] | 0.34 [-0.07, 0.76] | .028 |
| CADSS Amn | 0.94 (1.18) | 1.29 (1.19) | 2.03 | .076 | 0.29 [0.00, 0.74] | 0.58 (0.79) | 0.98 (1.12) | 2.10 | .051 | 0.40 [0.07, 0.76] | -0.11 [-0.55, 0.44] | .92 |
| CADSS Dep | 0.65 (1.01) | 2.74 (3.77) | 3.89 | <.001 | 0.74 [0.60, 1.15] | 0.81 (1.56) | 1.44 (2.23) | 1.84 | .088 | 0.32 [-0.01, 0.69] | 0.42 [0.05, 0.84] | .019 |
| CADSS Der | 2.68 (3.90) | 6.38 (6.62) | 4.37 | <.001 | 0.67 [0.40, 1.13] | 2.63 (3.80) | 4.05 (4.50) | 2.11 | .042 | 0.33 [0.04, 0.70] | 0.33 [-0.09, 0.82] | .039 |
| EDI | 6.49 (10.06) | 15.49 (21.20) | 2.84 | .007 | 0.53 [0.23, 0.92] | 4.22 (7.15) | 7.28 (14.25) | 1.45 | .16 | 0.27 [-0.06, 0.58] | 0.26 [-0.13, 0.71] | .11 |
| GASE | 2.78 (4.20) | 5.56 (6.36) | 4.02 | <.001 | 0.42 [0.23, 0.89] | 2.47 (3.92) | 4.02 (4.87) | 3.78 | <.001 | 0.35 [0.17, 0.63] | 0.07 [-0.25, 0.52] | .24 |

***Notes.*** 5D-ASC = *5-Dimensional Altered States of Consciousness Rating Scale*; 5D-ASC subscales: AUA = Auditory Alterations; DED = Dread of Ego Dissolution; OBN = Oceanic Boundlessness; VIR = Vigilance Reduction; VRS = Visionary Restructuralization; CADSS = *Clinician-Administered Dissociative States Scale*; CADSS Amn = CADSS-Amnesia subscale; CADSS-Dep = CADSS Depersonalization subscale; CADSS Der = CADSS-Derealization subscale; EDI = *Ego Dissolution Inventory*; GASE = *Generic Assessment of Side Effects*. *One participant removed from Blinded group due to a confidence rating of zero (equivalent to a random guess) in the placebo condition. All *p*- values are uncorrected permutation tests. Δ*g* reflects the difference between blinded and unblinded within-subject Hedges' *g* values; *p*-values are from permutation tests comparing participant-level placebo–control difference scores between groups. For comparisons of placebo–control effects by blinding status, all effects remained significant after Benjamini–Hochberg FDR correction adjusted *p* = .008).

*Compliance assessment*

Seventy-seven of 78 participants completed the five-point compliance scale. Participants reported low perceived compliance overall (*M* = 1.37, *SD* = 0.51). The majority of participants (65%; *n* = 49) indicated that all their responses were based solely on their experiences in the experiment, with an additional 35% (*n* = 27) reporting that most of their responses were based on their own experiences and only 1% (*n* = 1) reporting that their responses were equally based on their experiences and how they thought the researchers expected them to respond. No participants indicated that their responses were mostly or solely based on perceived experimental expectations. Compliance scores did not significantly differ between the blinded (*M* = 1.43, *SD* = 0.50) and unblinded (*M* = 1.33, *SD* = 0.53) participants, Z = 0.81, *p* = .51.

*Temporal reproduction*

Reproduction times (in ms) increased with target interval in both the control and placebo conditions (**Supplementary Figure 2**) and did not differ across conditions (see main text).

**Supplementary Figure 2.** *Temporal reproduction performance as a function of condition* (*N* = 74).

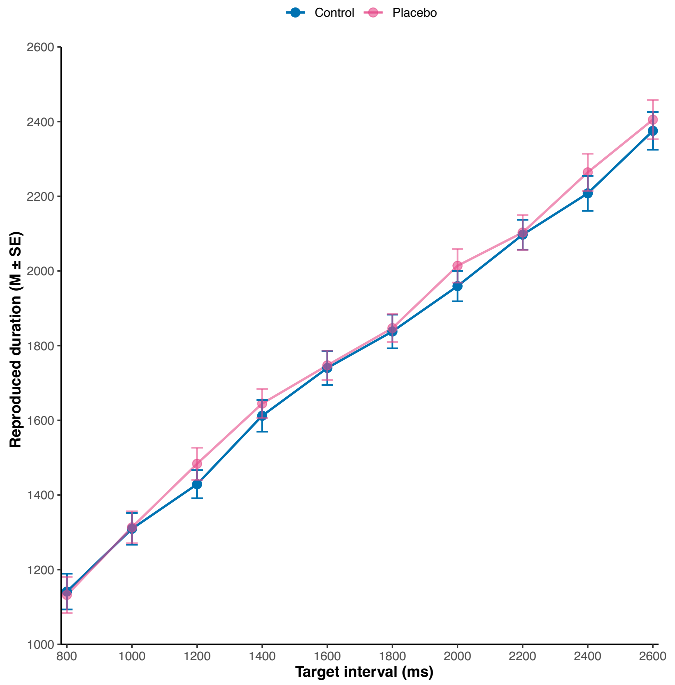

*Trait predictors of placebo and nocebo effects*

We conducted a series of regularized regression analyses to assess which individual difference variables predict the magnitude of placebo and nocebo effects (see Supplementary Table 5). Overall, models explained a small proportion of variance across outcomes. Cross-validated (CV) $R^2$ values were highly variable and often low (range = −.04 to .77), while in-sample $R^2$ estimates were consistently small (range = .00 to .15), indicating limited and potentially unstable predictive performance.

**Supplementary Table 5.** *Regularization regression results per outcome and optimal α penalty for each model (N = 78).*

| Outcome | Predictor | β [95% CI] | CV $R^2$ | $R^2$ | α |
|---|---|---|---|---|---|
| **5D-ASC** | | | .07 | .11 | 0 |
| | **BSS** | **0.22 [0.02, 0.72]** | | | |
| | **DES** | **0.02 [0.00, 0.03]** | | | |
| | **DES Abs** | **0.01 [0.00, 0.03]** | | | |
| | DES Amn | 0.00 [-0.01, 0.02] | | | |
| | DES DD | 0.01 [-0.00, 0.03] | | | |
| | ACE | 0.07[-0.05, 0.21] | | | |
| | SAS | 0.29 [-0.02, 0.81] | | | |
| | **GCS** | **0.03 [0.00, 0.12]** | | | |
| **5D-ASC AUA** | | | .06 | .08 | 0 |
| | BSS | 0.10 [-0.03, 0.39] | | | |
| | **DES** | **0.01 [0.00, 0.02]** | | | |
| | **DES Abs** | **0.00 [0.00, 0.02]** | | | |
| | DES Amn | 0.00 [-0.00, 0.01] | | | |
| | DES DD | 0.00 [-0.00, 0.02] | | | |
| | ACE | 0.04 [-0.01, 0.16] | | | |
| | SAS | 0.21 [-0.00, 0.58] | | | |
| | **GCS** | **0.02 [0.00, 0.04]** | | | |
| **5D-ASC DED** | | | .77 | .00 | 0.3 |
| | DES | 0.02 [0.00, 0.04] | | | |
| | DES Abs | 0.01 [0.00, 0.22] | | | |
| | DES Amn | 0.01 [-0.00, 0.14] | | | |
| **5D-ASC OBN** | | | -.29 | .15 | 0 |
| | BSS | 0.41 [-0.06, 1.43] | | | |
| | DES | **0.04 [0.00, 0.08]** | | | |
| | DES Abs | **0.03 [0.00, 0.07]** | | | |
| | DES Amn | -0.00 [-0.04, 0.02] | | | |
| | DES DD | **0.03 [0.00, 0.07]** | | | |
| | ACE | 0.19 [-0.06, 0.48] | | | |
| | SAS | 0.71 [-0.05, 1.82] | | | |
| | GCS | 0.03 [-0.01, 0.11] | | | |
| **5D-ASC VIR** | | | .12 | -.00 | 0 |
| | BSS | 0.01 [-0.00, 0.06] | | | |
| | DES | -0.00 [-0.00, 0.00] | | | |
| | DES Abs | 0.00 [-0.00, 0.00] | | | |
| | DES Amn | -0.00 [-0.00, 0.00] | | | |
| | DES DD | -0.00 [-0.00, 0.00] | | | |
| | ACE | -0.01 [-0.02, 0.00] | | | |

| Outcome | Predictor | β [95% CI] | CV $R^2$ | $R^2$ | α |
|---|---|---|---|---|---|
| | SAS | 0.00 [-0.01, 0.06] | | | |
| | GCS | 0.00 [-0.002, 0.01] | | | |
| **5D-ASC VRS** | | | -.29 | .23 | 1 |
| | BSS | 0.48 [0.00, 2.40] | | | |
| | DES Abs | 0.06 [0.00, 0.13] | | | |
| | DES DD | 0.04 [0.00, 0.15] | | | |
| | ACE | 0.40 [0.00, 1.07] | | | |
| **CADSS** | | | .03 | .15 | 1 |
| | DES Abs | 0.09 [0.00, 0.14] | | | |
| | SAS | -1.22 [-2.90, 0.00] | | | |
| | **GCS** | **0.36 [0.06, 1.05]** | | | |
| **CADSS Amn** | | | 0.05 | .00 | 0 |
| | BSS | -0.00 [-0.00, 0.01] | | | |
| | DES | 0.00 [0.00, 0.00] | | | |
| | DES Abs | 0.00 [0.00, 0.00] | | | |
| | DES Amn | 0.00 [-0.00, 0.00] | | | |
| | DES DD | 0.00 [-0.00, 0.00] | | | |
| | ACE | 0.00 [0.00, 0.00] | | | |
| | SAS | -0.00 [-0.02, 0.01] | | | |
| | GCS | 0.00[-0.00, 0.00] | | | |
| **CADSS Dep** | | | .11 | .13 | 0.6 |
| | DES | 0.01 [0.00, 0.05] | | | |
| | DES Abs | 0.01 [0.00, 0.04] | | | |
| | DES DD | 0.01 [0.00, 0.04] | | | |
| | GCS | 0.12 [0.00, 0.35] | | | |
| **CADSS Der** | | | -.04 | .12 | 0.4 |
| | DES | 0.00 [0.00, 0.03] | | | |
| | DES Abs | 0.04 [0.00, 0.07] | | | |
| | ACE | 0.03 [-0.37, 0.38] | | | |
| | SAS | -0.97 [-1.84, 0.00] | | | |
| | **GCS** | **0.19 [0.03, 0.57]** | | | |
| **EDI** | | | -.06 | .12 | 0.2 |
| | DES | 0.04 [0.00, 0.15] | | | |
| | **DES Abs** | **0.14 [0.01, 0.31]** | | | |
| | DES Amn | -0.19 [-0.29, 0.08] | | | |
| | DES DD | 0.08 [0.00, 0.30] | | | |
| | GCS | 0.37 [0.00, 1.37] | | | |
| **GASE** | | | .05 | .13 | 0.9 |
| | DES | 0.02 [0.00, 0.07] | | | |
| | DES Abs | 0.02 [0.00, 0.07] | | | |
| | DES Amn | 0.01 [0.00, 0.06] | | | |

*Notes.* Bolded values reflect significant effects in which 95% CIs do not cross zero; CV = cross-validated; α values represent the mixing parameter in elastic net models where 0 corresponds to ridge regression, 1 corresponds to lasso regression, and intermediate values to a combination of both (elastic net).

#### *Moderators of the nocebo effect*

We prespecified four simple moderation and one moderated-moderation analyses to assess associations between childhood trauma (ACE), dissociation (DES), absorption (DES Abs), depersonalization/derealization (DES DD), and somatosensory amplification (SAS) on the magnitude of the nocebo effect (placebo-control GASE scores; **Supplementary Table 6**). Consistent with our pre-registered hypotheses, DES, DES-DD, DES-Abs emerged as significant independent predictors of the magnitude of the nocebo effect. In contrast, ACE and SAS did not significantly predict the nocebo effect. Across all models, there were no significant one- or two-way interaction effects. Model comparison based on Bayesian Information Criterion (BIC) indicated that the ACE × DES-Abs model provided the best fit to the data (BIC = 437.23). These results suggest that different facets of dissociation independently predict the magnitude of the nocebo effect, but that childhood trauma and somatosensory amplification do not moderate these associations, nor independently predict outcomes, in the present sample.

**Supplementary Table 6.** *Inferential statistics for simple moderation and moderated-moderation models predicting the nocebo effect (N = 77*)*

| Model | Predictor | *b* | SE | *t* | *p* | $R^2$ | BIC |
|---|---|---|---|---|---|---|---|
| ACE × DES | $F(3, 73) = 4.01, p = .011$ | | | | | .14 | 440.56 |
| | **DES** | **0.09** | **0.03** | **3.33** | **<.001** | | |
| | ACE | 0.03 | 0.21 | 0.14 | .88 | | |
| | ACE × DES | -0.01 | 0.01 | -0.40 | .69 | | |
| ACE × DES Abs | $F(3, 73) = 4.53, p = .006$ | | | | | .16 | 437.23 |
| | **DES Abs** | **0.07** | **0.02** | **3.55** | **<.001** | | |
| | ACE | 0.09 | 0.17 | 0.53 | .60 | | |
| | ACE × DES Abs | -0.01 | 0.01 | -0.57 | .57 | | |
| ACE × DD | $F(3, 73) = 1.64, p = .18$ | | | | | .06 | 447.65 |
| | DD | 0.05 | 0.03 | 1.94 | .057 | | |
| | ACE | 0.09 | 0.21 | 0.43 | .66 | | |
| | ACE × DD | -0.01 | 0.02 | -0.84 | .40 | | |
| ACE × SAS | $F(3, 73) = 0.45, p = .717$ | | | | | .02 | 452.34 |
| | ACE | 0.15 | 0.18 | 0.82 | .41 | | |
| | SAS | 0.24 | 0.58 | 0.42 | .68 | | |
| | ACE × SAS | -0.17 | 0.27 | -0.63 | .53 | | |
| ACE × DES × SAS | $F(7, 69) = 2.22, p = .043$ | | | | | .18 | 443.87 |
| | **DES** | **0.10** | **0.03** | **3.35** | **<.001** | | |
| | ACE | -0.00 | 0.16 | -0.01 | .99 | | |
| | SAS | -0.37 | 0.61 | -0.61 | .54 | | |
| | DES × ACE | 0.01 | 0.03 | 0.37 | .71 | | |
| | DES × SAS | -0.09 | 0.06 | -1.55 | .12 | | |
| | ACE × SAS | -0.05 | 0.29 | -0.17 | .86 | | |
| | DES × ACE × SAS | -0.00 | 0.03 | -0.08 | .93 | | |

*Notes.* *One participant removed due to missing response on ACE; ACE = Adverse Childhood Experiences; DES = Dissociative Experiences Scale; SAS = Somatosensory Amplification Scale; DD = Depersonalisation/Derealisation.

**References**

Barnes, K., Faasse, K., Geers, A. L., Helfer, S. G., Sharpe, L., Colloca, L., & Colagiuri, B. (2019). Can positive framing reduce nocebo side effects? Current evidence and recommendation for future research. *Front Pharmacol*, *10*, 167.

Barsky, A. J., Wyshak, G., & Klerman, G. L. (1990). The somatosensory amplification scale and its relationship to hypochondriasis. *J Psychiatr Res*, *24*(4), 323-334.

Bowers, K. S. (1981). Do the stanford scales tap the "classic suggestion effect"? *Int J Clin Exp Hypn*, *29*(1), 42-53.

Bowers, K. S. (1998). Waterloo-stanford group scale of hypnotic susceptibility, form c: Manual and response booklet. *Int J Clin Exp Hypn*, *46*(3), 250-268.

Bowers, P., Laurence, J. R., & Hart, D. (1988). The experience of hypnotic suggestions. *Int J Clin Exp Hypn*, *36*(4), 336-349.

Brake, B., Wieder, L., Hughes, N., Lalinde, I. S., Marr, D., Geagea, D., Pick, S., Reinders, A., Kamboj, S. K., Thompson, T., & Terhune, D. B. (2025). The induction of dissociative states: A meta-analysis. *Biol Psychiatry Glob Open Sci*, *5*(4), 100521.

Bremner, J. D., Krystal, J. H., Putnam, F. W., Southwick, S. M., Marmar, C., Charney, D. S., & Mazure, C. M. (1998). Measurement of dissociative states with the clinician-administered dissociative states scale (cadss). *J Trauma Stress*, *11*(1), 125-136.

Carlson, E. B., & Putnam, F. W. (1993). An update on the dissociative experiences scale. *Dissociation: Progress in the Dissociative Disorders*, *6*(1), 16-27.

Das, R. K., Tamman, A., Nikolova, V., Freeman, T. P., Bisby, J. A., Lazzarino, A. I., & Kamboj, S. K. (2016). Nitrous oxide speeds the reduction of distressing intrusive memories in an experimental model of psychological trauma. *Psychol Med*, *46*(8), 1749-1759.

Dittrich, A. (1998). The standardized psychometric assessment of altered states of consciousness (ascs) in humans. *Pharmacopsychiatry*, *31 Suppl 2*, 80-84.

Doering, B. K., Szécsi, J., Bárdos, G., & Köteles, F. (2016). Somatosensory amplification is a predictor of self-reported side effects in the treatment of primary hypertension: A pilot study. *Int J Behav Med*, *23*(3), 327-332.

Earleywine, M., Mian, M. N., Altman, B. R., & De Leo, J. A. (2022). Expectancies for cannabis-induced emotional breakthrough, mystical experiences and changes in dysfunctional attitudes: Perceptions of the potential for cannabis-assisted psychotherapy for depression. *Cannabis*, *5*(2), 16-27.

Erceg-Hurn, D. M., & Mirosevich, V. M. (2008). Modern robust statistical methods: An easy way to maximize the accuracy and power of your research. *Am Psychol*, *63*(7), 591-601.

Felitti, V. J., Anda, R. F., Nordenberg, D., Williamson, D. F., Spitz, A. M., Edwards, V., Koss, M. P., & Marks, J. S. (1998). Relationship of childhood abuse and household dysfunction to many of the leading causes of death in adults. The adverse childhood experiences (ace) study. *Am J Prev Med*, *14*(4), 245-258.

Gudjonsson, G. H. (1989). Compliance in an interrogative situation: A new scale. *Personality and Individual Differences*, *10*(5), 535-540.

Jensen, M. E., Stenbæk, D. S., Juul, T. S., Fisher, P. M., Ekstrøm, C. T., Knudsen, G. M., & Fink-Jensen, A. (2022). Psilocybin-assisted therapy for reducing alcohol intake in patients with alcohol use disorder: Protocol for a randomised, double-blinded, placebo-controlled 12-week clinical trial (the quantum trip trial). *BMJ Open*, *12*(10), e066019.

Kamboj, S. K., Gong, A. T., Sim, Z., Rashid, A. A., Baba, A., Iskandar, G., Das, R. K., & Curran, H. V. (2020). Reduction in the occurrence of distressing involuntary memories following propranolol or hydrocortisone in healthy women. *Psychol Med*, *50*(7), 1148-1155.

Morgan, C. J., Mofeez, A., Brandner, B., Bromley, L., & Curran, H. V. (2004). Acute effects of ketamine on memory systems and psychotic symptoms in healthy volunteers. *Neuropsychopharmacology*, *29*(1), 208-218.

Nash, R., Saraiva, R., & Hope, L. (2022). Who doesn’t believe their memories? Development and validation of a new memory distrust scale. *Journal of Applied Research in Memory and Cognition*, *12*.

Niciu, M. J., Shovestul, B. J., Jaso, B. A., Farmer, C., Luckenbaugh, D. A., Brutsche, N. E., Park, L. T., Ballard, E. D., & Zarate, C. A., Jr. (2018). Features of dissociation differentially predict antidepressant response to ketamine in treatment-resistant depression. *J Affect Disord*, *232*, 310-315.

Nour, M. M., Evans, L., Nutt, D., & Carhart-Harris, R. L. (2016). Ego-dissolution and psychedelics: Validation of the ego-dissolution inventory (edi). *Front Hum Neurosci*, *10*, 269.

Piazza, G. G., Iskandar, G., Hennessy, V., Zhao, H., Walsh, K., McDonnell, J., Terhune, D. B., Das, R. K., & Kamboj, S. K. (2022). Pharmacological modelling of dissociation and psychosis: An evaluation of the clinician administered dissociative states scale and psychotomimetic states inventory during nitrous oxide ('laughing gas')-induced anomalous states. *Psychopharmacology (Berl)*, *239*(7), 2317-2329.

Prugger, J., Derdiyok, E., Dinkelacker, J., Costines, C., & Schmidt, T. T. (2022). The altered states database: Psychometric data from a systematic literature review. *Scientific Data*, *9*(1), 720.

Rief, W., Barsky, A. J., Glombiewski, J. A., Nestoriuc, Y., Glaesmer, H., & Braehler, E. (2011). Assessing general side effects in clinical trials: Reference data from the general population. *Pharmacoepidemiol Drug Saf*, *20*(4), 405-415.

Shor, R. E., & Orne, E. C. (1962). *Harvard group scale of hypnotic susceptibility, form a.* Consulting Psychologists Press.

Skovbjerg, S., Zachariae, R., Rasmussen, A., Johansen, J. D., & Elberling, J. (2010). Attention to bodily sensations and symptom perception in individuals with idiopathic environmental intolerance. *Environ Health Prev Med*, *15*(3), 141-150.

Studerus, E., Gamma, A., & Vollenweider, F. X. (2010). Psychometric evaluation of the altered states of consciousness rating scale (oav). *PLoS One*, *5*(8), e12412.

Szigeti, B., Nutt, D., Carhart-Harris, R., & Erritzoe, D. (2023). The difference between 'placebo group' and 'placebo control': A case study in psychedelic microdosing. *Scientific Reports*, *13*(1), 12107.

Weitzenhoffer, A. M., & Hilgard, E. R. . (1962). *Stanford hypnotic susceptibility scale: Form c.* Consulting Psychologists Press.

Wieder, L., & Terhune, D. B. (2019). Trauma and anxious attachment influence the relationship between suggestibility and dissociation: A moderated-moderation analysis. *Cognitive Neuropsychiatry*, *24*(3), 191-207.

Winkler, J., Kanouse, D., & Ware, J. (1982). Controlling for acquiescence response set in scale development. *Journal of Applied Psychology*, *67*, 555-561.
